# Children with SARS-CoV-2 in the National COVID Cohort Collaborative (N3C)

**DOI:** 10.1101/2021.07.19.21260767

**Authors:** Blake Martin, Peter E. DeWitt, Seth Russell, Adit Anand, Katie R. Bradwell, Carolyn Bremer, Davera Gabriel, Andrew T. Girvin, Janos G. Hajagos, Julie A. McMurry, Andrew J. Neumann, Emily R. Pfaff, Anita Walden, Jacob T. Wooldridge, Yun Jae Yoo, Joel Saltz, Ken R. Gersing, Christopher G. Chute, Melissa A. Haendel, Richard Moffitt, Tellen D. Bennett

**Affiliations:** Section of Critical Care Medicine, Department of Pediatrics, University of Colorado School of Medicine, University of Colorado, Aurora, CO, USA; Section of Informatics and Data Science, Department of Pediatrics, University of Colorado School of Medicine, University of Colorado, Aurora, CO, USA; Department of Biomedical Informatics, Stony Brook University, Stony Brook, NY, USA; Palantir Technologies, Denver, CO, USA; Johns Hopkins University School of Medicine, Baltimore, MD, USA; Translational and Integrative Sciences Center, University of Colorado, Aurora, CO, USA; Center for Health AI, University of Colorado, Aurora, CO, USA; North Carolina Translational and Clinical Sciences Institute (NC TraCS), University of North Carolina at Chapel Hill, Chapel Hill, NC, USA; National Center for Advancing Translational Sciences, National Institutes of Health, Bethesda, MD, USA; Schools of Public Health, and Nursing, Johns Hopkins University, Baltimore, MD, USA

**Author notes:** **Corresponding Author:** Blake Martin, MD, Department of Pediatrics, University of Colorado School of Medicine, 13121 E. 17th Avenue, MS8414, Aurora, CO 80045.

## Abstract

**Importance:** SARS-CoV-2

**Objective:** To determine the characteristics, changes over time, outcomes, and severity risk factors of SARS-CoV-2 affected children within the National COVID Cohort Collaborative (N3C)

**Design:** Prospective cohort study of patient encounters with end dates before May 27th, 2021.

**Setting:** 45 N3C institutions

**Participants:** Children <19-years-old at initial SARS-CoV-2 testing

**Main Outcomes and Measures:** Case incidence and severity over time, demographic and comorbidity severity risk factors, vital sign and laboratory trajectories, clinical outcomes, and acute COVID-19 vs MIS-C contrasts for children infected with SARS-CoV-2.

**Results:** 728,047 children in the N3C were tested for SARS-CoV-2; of these, 91,865 (12.6%) were positive. Among the 5,213 (6%) hospitalized children, 685 (13%) met criteria for severe disease: mechanical ventilation (7%), vasopressor/inotropic support (7%), ECMO (0.6%), or death/discharge to hospice (1.1%). Male gender, African American race, older age, and several pediatric complex chronic condition (PCCC) subcategories were associated with higher clinical severity (p*≤*0.05). Vital signs (all p≤0.002) and many laboratory tests from the first day of hospitalization were predictive of peak disease severity. Children with severe (vs moderate) disease were more likely to receive antimicrobials (71% vs 32%, p<0.001) and immunomodulatory medications (53% vs 16%, p<0.001).

Compared to those with acute COVID-19, children with MIS-C were more likely to be male, Black/African American, 1-to-12-years-old, and less likely to have asthma, diabetes, or a PCCC (p*<*0.04). MIS-C cases demonstrated a more inflammatory laboratory profile and more severe clinical phenotype with higher rates of invasive ventilation (12% vs 6%) and need for vasoactive-inotropic support (31% vs 6%) compared to acute COVID-19 cases, respectively (p<0.03).

**Conclusions:** In the largest U.S. SARS-CoV-2-positive pediatric cohort to date, we observed differences in demographics, pre-existing comorbidities, and initial vital sign and laboratory test values between severity subgroups. Taken together, these results suggest that early identification of children likely to progress to severe disease could be achieved using readily available data elements from the day of admission. Further work is needed to translate this knowledge into improved outcomes.

## Introduction

As of June, 2021, severe acute respiratory syndrome associated with coronavirus-2 (SARS-CoV-2) has infected more than 177 million people and caused more than 3.8 million deaths worldwide ^1^. SARS-CoV-2 can cause coronavirus disease of 2019 (COVID-19), a condition characterized by pneumonia, hypoxemic respiratory failure, thrombosis, cardiac and renal dysfunction, hyperinflammation, and substantial mortality and morbidity. Though children often experience milder illness^2-4^, SARS-CoV-2 can cause severe pediatric disease in the form of both acute COVID-19 and multisystem inflammatory syndrome in children (MIS-C). Both acute COVID-19 and MIS-C can cause significant morbidity and mortality^5-10^. MIS-C is a hyperinflammatory condition thought to represent a post-infectious complication of SARS-CoV-2 infection^5,11,12^. It is characterized by cardiovascular, respiratory, neurologic, gastrointestinal, and mucocutaneous manifestations and organ dysfunction. Thus far, over 1,700 cases of MIS-C have been reported to the U.S. Centers for Disease Control and Prevention, with over half requiring ICU admission and greater than one-third experiencing shock^13^.

Research in pediatric COVID-19 and MIS-C has been slowed by the lack of large, multi-institutional datasets of affected children. Investigators in Europe^7^ and the U.S.^10,13,14^ have reported multi-center studies, but analysis of individual patient vital sign and laboratory data in those cohorts was absent. An extensive, granular, representative clinical dataset is needed to improve our understanding of the presentation, risk factors, and severity signals of pediatric COVID-19 and MIS-C.

The National COVID Cohort Collaborative (N3C) was formed to improve understanding of SARS-CoV-2 infections and clinical outcomes via a novel approach to data sharing and analytics. The N3C^15^ is comprised of members from the NIH Clinical and Translational Science Awards Program, its Center for Data to Health, the IDeA Centers for Translational Research^16^, and several EHR-based research networks^17^. We have previously reported details regarding N3C structure, data ingestion and integration, the number of N3C sites using each common data model (CDM), and the patient variables supported by each CDM^15,17^. See supplemental methods for information on N3C patient inclusion criteria and available data, and details regarding N3C funding sources.

Our objective was to provide a detailed clinical characterization of the largest cohort of U.S. pediatric SARS-CoV-2 positive cases to date. We hypothesized that we would be able to 1) identify risk factors for higher severity disease among hospitalized children, 2) evaluate rates of SARS-CoV-2 positive cases and hospitalizations over time, 3) visualize changes in SARS-CoV-2 medication treatment regimens over time, 4) compare trends in vital sign and laboratory markers between hospitalized children in different clinical severity subgroups, and 5) identify differences in clinical risk factors and outcomes between children with acute COVID-19 versus MIS-C.

## Methods & Study Design

### Cohort definition

We performed a retrospective analysis of all children <19-years-old at first SARS-CoV-2 testing at the 45 N3C sites whose data 1) completed harmonization and integration, 2) were released for analysis, and 3) included the necessary death and mechanical ventilation information. Patients were included if their index encounter (see supplemental methods) ended before May 27th, 2021.

Children were considered to have SARS-CoV-2 if they had a documented positive test by polymerase chain reaction (PCR), antigen (Ag), or antibody (Ab). Children were considered to have MIS-C if they were 1) hospitalized with a positive SARS-CoV-2 test and 2) assigned either of two recommended ICD-10 diagnosis codes for MIS-C; during the early phase of the pandemic, M35.8^18^, or the more specific M35.81 introduced October 1st, 2020 and effective January 1st, 2021^19^.

We described the geographic location of pediatric N3C patients including the incidence of positive cases by month with comparison to the adult N3C cohort. We evaluated changes in pediatric age distribution and maximum clinical severity over time (see below for clinical severity definition). We determined the proportion of hospitalized children who received different antimicrobial and immunomodulatory medications by month and provided corresponding adult trends for comparison. Lastly, we compared the clinical characteristics, outcomes, and laboratory test profiles between children with MIS-C vs those with acute COVID-19 (children with a positive SARS-CoV-2 PCR or Ag test but no MIS-C diagnosis code nor positive antibody test result).

### Hospital Index Encounter and Clinical Severity

We identified a single clinical encounter for each laboratory-confirmed SARS-CoV-2 positive child by selecting the encounter demonstrating the maximum Clinical Progression Scale (CPS) score (created by the World Health Organization [WHO] for COVID-19 clinical research^17,20^) (see Supplement). Clinical severity categories included mild (outpatient or emergency department visits only), moderate (hospitalized without need for invasive ventilation, vasopressor / inotropic medications, or ECMO), and severe (hospitalized and requiring invasive ventilation, vasopressors / inotropes, or ECMO, or death, or discharge to hospice).

### Variable Definition and Statistical Methods

For each clinical concept (e.g., laboratory measure, vital sign, medication, or preselected comorbidity) we defined or identified existing established concept sets in the Observational Medical Outcomes Partnership (OMOP)^21^ CDM. We used established concept sets to identify children with the preselected comorbidities of asthma, diabetes mellitus (types I and II), and obesity given prior reports of more severe disease among such patients^8,10^. We identified children with chronic medical conditions for disease-severity risk analysis using the pediatric complex chronic conditions (PCCC) algorithm via adaptation of our prior R implementation^22,23^. We validated our newly defined concept sets with input from informatics and clinical subject matter experts. All concept sets and analytic pipelines are fully reproducible and will be made publicly available.

We tested demographic differences in the relative proportions of moderate and severe patients via logistic regression models. We used chi-square deviance tests to evaluate changes in relative proportions of predefined pediatric age ranges over time and to evaluate changes in the proportion of moderate and severe cases over time. We evaluated differences in initial vital sign and lab values between moderate and severe subgroups using generalized estimating equations. We assessed for significance in change over time for each vital sign or lab result during a patient’s hospitalization; we did this using a linear model to average over the first week of values and then used the model results to estimate the difference between hospital day 0 and day 7 for each severity subgroup. See Supplemental Methods for additional information including software packages used.

## Results

### Study Cohort Demographics and Comorbidities

The N3C dataset released May 27th, 2021 contains 728,047 pediatric patients, of which 91,865 (13%) had a positive SARS-CoV-2 test (PCR/Ag, or Ab) (Table 1, Supplemental Table 1). The 45 hospital systems from which these data were obtained were located in several US geographic subregions (defined using the US Census Bureau divisions) and were most concentrated in the South Atlantic and East North Central regions (Figure 1). Figures 1c and 1d demonstrate the evolution of monthly pediatric SARS-CoV-2 cases by geographic region and overall for each month during the study period, which closely mimics the N3C adult case incidence as shown. The incidence of positive SARS-CoV-2 tests peaked in November, 2020 with peak hospitalization incidence in December, 2020 for adults and children (Figure 2b).

**Table 1:**
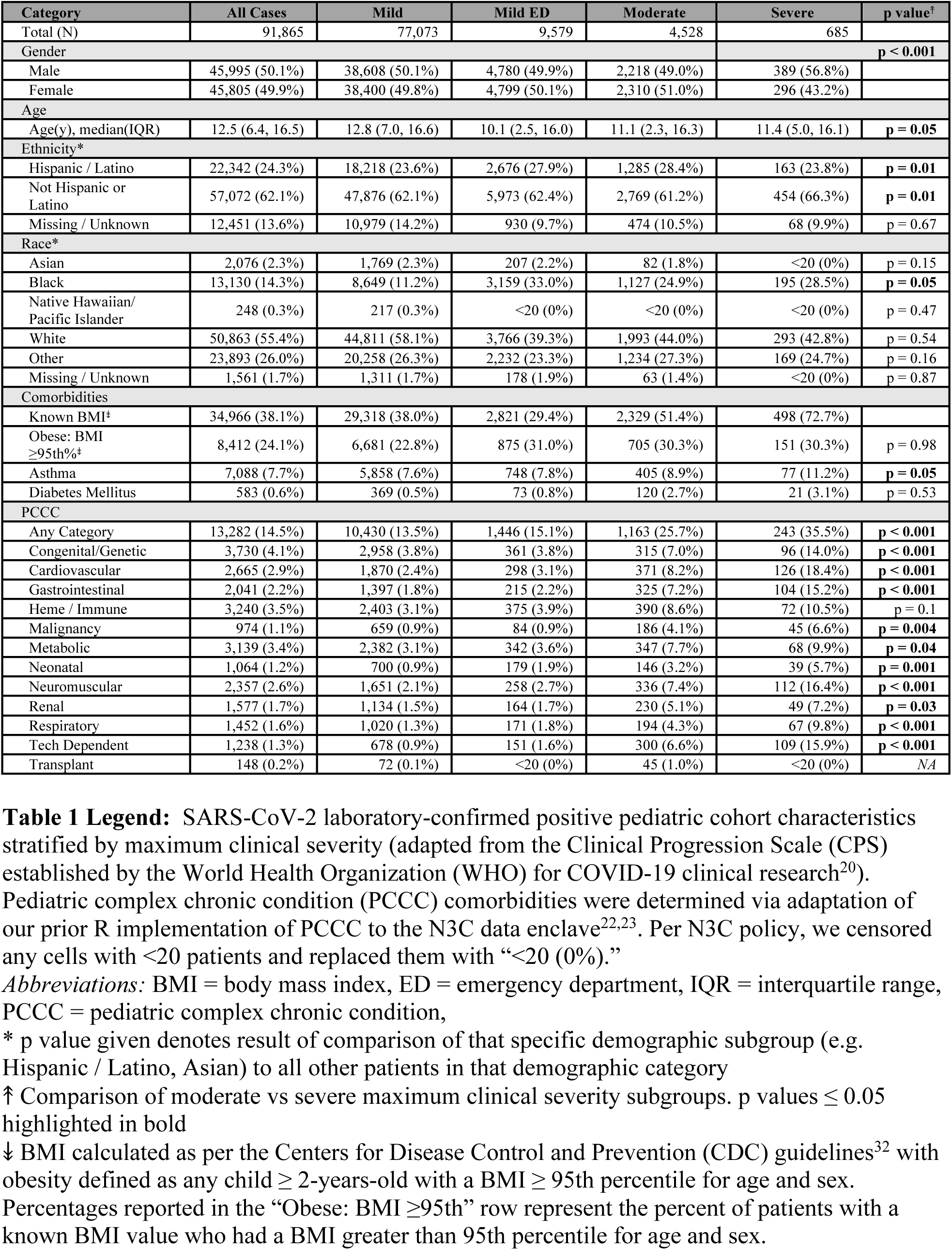
Demographics and Comorbidities for SARS-CoV-2 Positive Children.

**Figure 1:**
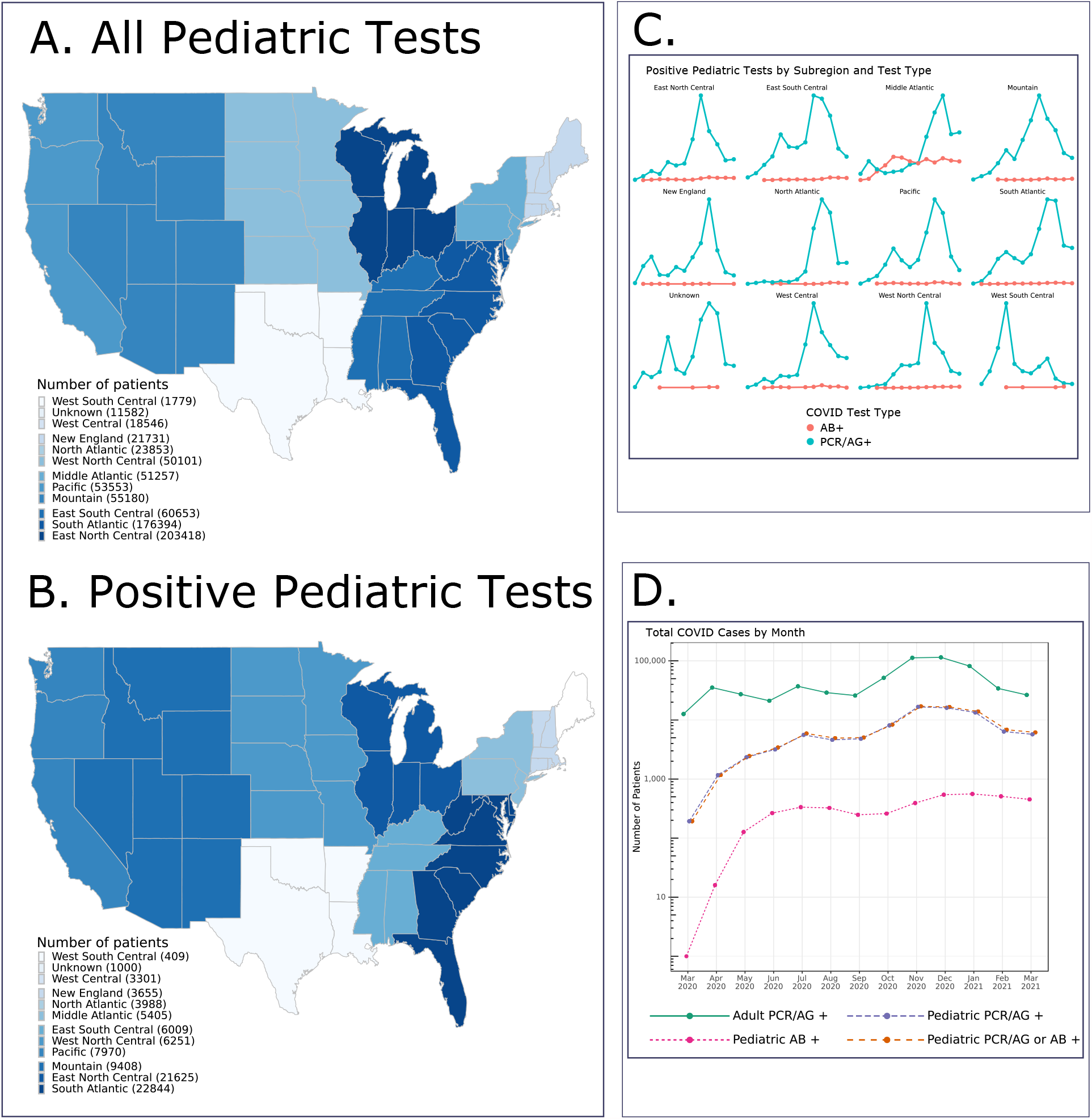
Geographic distribution and case incidence over time for SARS-CoV-2 positive patients. **Figure 1a** shows the geographic distribution of all pediatric N3C patients (N = 728,047). **Figure 1b** shows the geographic distribution of the positive pediatric cases only (N = 91,865). **Figure 1c** shows the monthly trends for positive pediatric SARS-CoV-2 testing by subregion and test type. **Figure 1d** shows the monthly trends for SARS-CoV-2 positive children by test type along with overall adult positive PCR/Ag cases (N = 609,734) for comparison.

**Figure 2:**
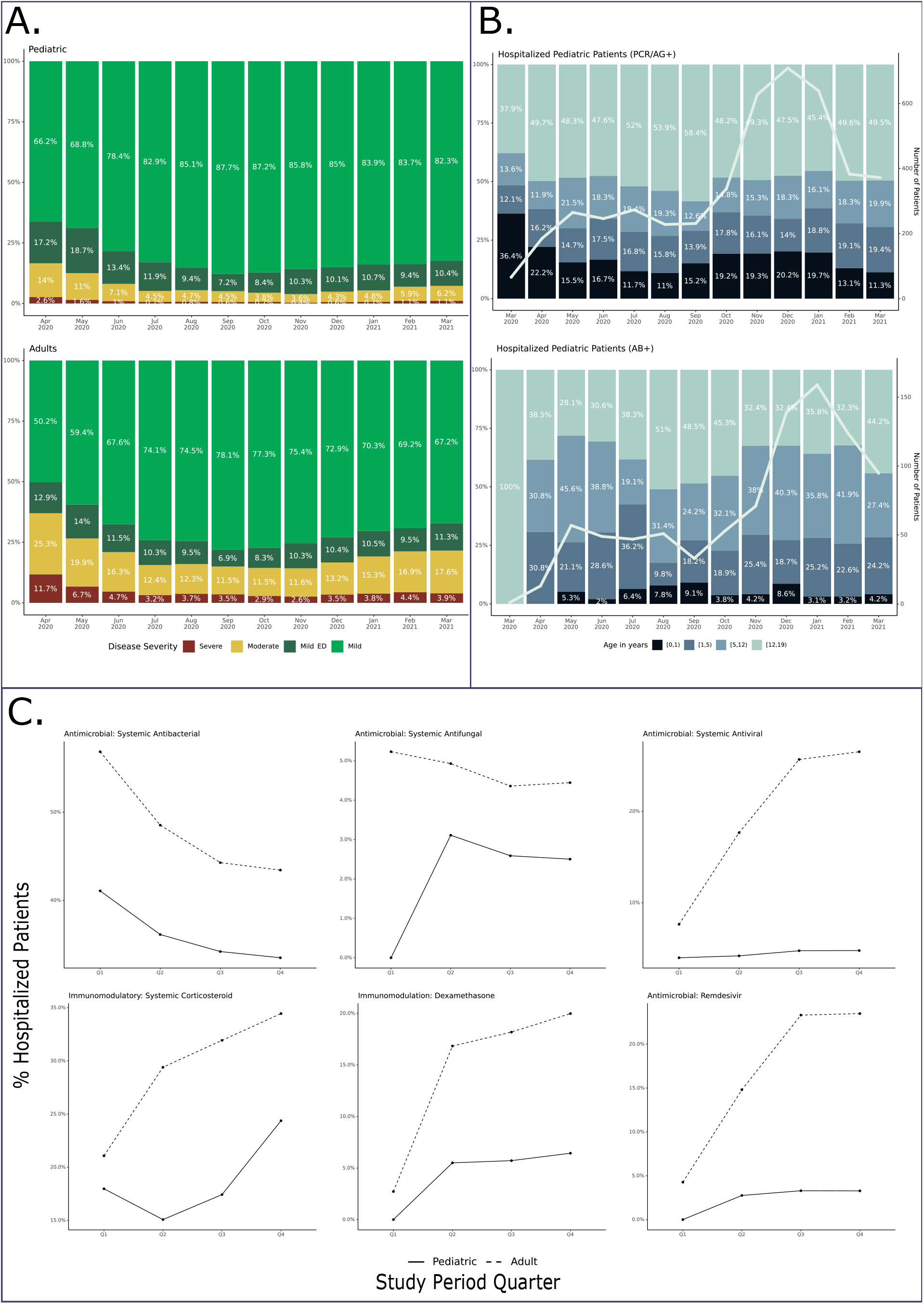
Age, maximum clinical severity, and antimicrobial and immunomodulatory medication use over time for SARS-CoV-2 positive children. **Figure 2a** illustrates changes in the distribution of maximum clinical severity (by WHO CPS score) by month during the study period compared to N3C positive adults. Red = hospital mortality, discharge to hospice, or invasive ventilation, vasoactive-inotropic support, or ECMO. Yellow = Hospitalized without any of those. Dark Green = Emergency Department visit. Light Green = Outpatient visit. March, 2020 censored given <20 pediatric patients in the severe subgroup. **Figure 2b** shows the age category distribution of infected children by month during the study period stratified by test type (PCR/Ag+ with negative or no Ab testing vs Ab+ regardless of PCR/Ag testing results). The trendline demonstrates the monthly positive test incidence. **Figure 1c** shows the evolution in use of selected antimicrobial and immunomodulatory medications by quarter (Apr 2020 - Mar 2021) among hospitalized children with SARS-CoV-2 compared to hospitalized N3C adult cases: Q1 = Apr 2020 - Jun 2020, Q2 = Jul 2020 - Sep 2020, Q3 = Oct 2020 - Dec 2020, and Q4 = Jan 2021 - Mar 2021

The demographics of the SARS-CoV-2 positive pediatric cohort are shown in Table 1, stratified by maximum clinical severity. The cohort is a diverse group representative of many segments of the US pediatric population. Compared to the moderate severity subgroup patients, the severe subgroup patients were more likely to be older, male, African-American, and have asthma or a PCCC (except the hematologic/immunologic category)(p *≤* 0.05 for all).

### Clinical Course and Illness Severity

Overall, 5,213 (5.7%) children were hospitalized. Of these, 355 (6.8%) required mechanical ventilation, and 57 (1.1%) died. The 1.1% mortality rate is consistent with two pediatric reports (0.9% and 2%)^10,14^ and lower than the 11.6% rate reported for hospitalized adults in the N3C cohort^17^. We and others have reported a decrease in adult COVID-19 severity as the pandemic has progressed.^17,24^ We also observed a statistically significant decrease in the proportion of moderate and severe pediatric cases over the study period (p < 0.001) (Figure 2a).

### Age Distribution

The age distributions for SARS-CoV-2 positive children are illustrated in Figure 2b. Most patients were 12-17 years of age. The age distribution had a statistically significant change (p < 0.001) during the study period increasing in the 1-5 and 5-12 age groups. Among Ab+ (positive antibody test regardless of PCR/Ag testing) hospitalized children, we observed a larger proportion of 1-5 and 5-12 year old patients with a later peak incidence in January, 2021. As antibody testing may be a surrogate for MIS-C evaluation, the timing of this peak is consistent with prior studies demonstrating maximal MIS-C risk in the 2-to-5 weeks after initial SARS-CoV-2 infection^13^.

### Treatments

Use of antimicrobial and immunomodulatory medications changed considerably during the study period (Figure 2c). As with N3C adults^17^, a higher proportion of children from the severe subgroup received antimicrobials compared to the moderate subgroup (71% vs 32%), with antibacterials (68% vs 31%) and antivirals (14% vs 3%) specifically being more common (p < 0.001 for all)(Supplemental Table 2). We observed a decrease in azithromycin use over time in both pediatric and adult patients, presumably related to accumulating evidence regarding lack of efficacy.

Immunomodulatory medication use was also more common among severely ill children compared to the moderately ill (53% vs 16%, p < 0.001 overall and for each medication). Systemic corticosteroid (50% vs 15%, p < 0.001) and anakinra (9% vs 0.8%, p < 0.001) use was markedly higher in the severe subgroup than in the moderate subgroup.

Of the 5,213 hospitalized children, 1,931 (37%) received at least one antimicrobial and 1,071 (20%) received at least one immunomodulatory medication with 1,004 (19%) specifically receiving a systemic corticosteroid. Antivirals with potential activity against SARS-CoV-2 (remdesivir) were administered to 148 (3%) of hospitalized children with higher frequency use among the severe subgroup compared to moderate (9% vs 2%, p < 0.001). Of note, the FDA’s Emergency Use Authorization for remdesivir^25^ occurred on May, 1^st^, 2020, and the trial indicating survival benefit from dexamethasone^26^ as well as a landmark publication describing MIS-C^5^ were both published in July, 2020.

### Vital Sign and Laboratory Measurements

Compared to the moderate severity subgroup, children in the severe subgroup had significantly more abnormal initial values for many vital signs including systolic and diastolic blood pressure (lower), SpO2 (lower), heart rate (higher), and respiratory rate (higher) (p ≤ 0.002 for all). We saw improvement (values becoming more normal) in most vital signs from day 0 to day 7 for both subgroups (Figure 3)(Supplemental Table 3).

**Figure 3:**
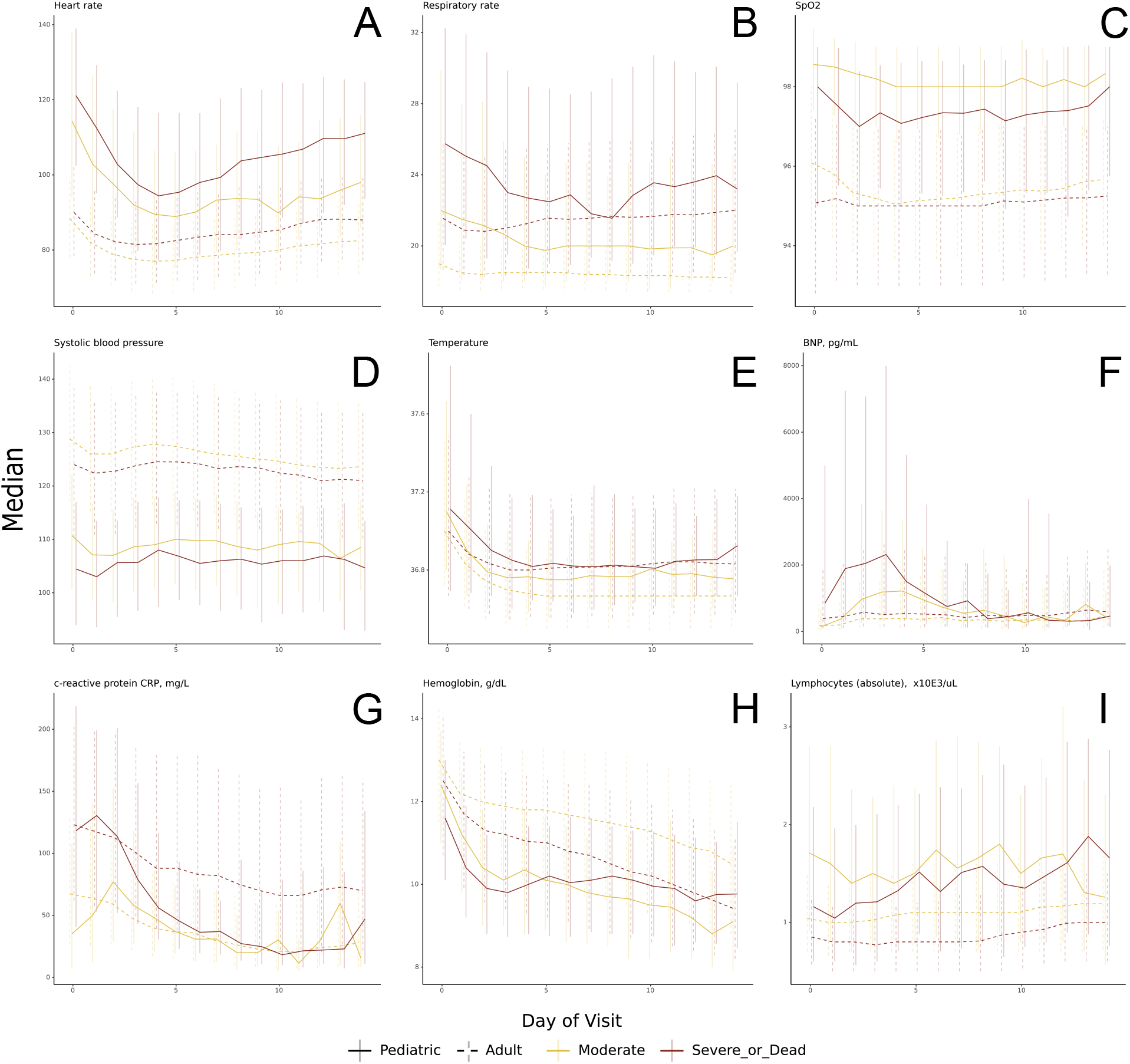
Vital sign and laboratory value trajectories. Trajectories of selected vital sign (a-e) and laboratory (f-i) median values by day of hospitalization during pediatric hospital encounters as compared to N3C adult values, stratified my maximum clinical severity

Children who eventually experienced the highest maximum clinical severity also had many laboratory tests whose initial values were more abnormal than in the moderate severity subgroup (Figure 3, Supplemental Table 4). Specifically, initial median values were more abnormal in the severe subgroup for ALT and AST (higher), albumin (lower), BNP (higher), creatinine (higher), D-dimer (higher), ferritin (higher), CRP (higher), and total white blood cell count (higher)(p < 0.05 for all). Comparing hospital day 0 to day 7, we observed a trend toward normalization for most laboratory tests within both subgroups. Exceptions include more abnormal values for albumin and hemoglobin (lower) in moderate and severe subgroups and erythrocyte sedimentation rate (higher) within the moderate subgroup (p < 0.05 for all). See Supplement Table 5 for proportions of children with each lab test available. The improving trajectories for most vital sign and laboratory values likely reflect the low mortality rate (and high recovery rate) of infected children.

### Acute COVID vs MIS-C

We used specific ICD-10 codes to identify 498 patients with MIS-C, of which 439 (88%) were hospitalized and 153 (31%) met criteria for severe disease. The demographics, lab results, and clinical outcomes for children with MIS-C versus acute COVID-19 (positive PCR/Ag test but no Ab+ test nor MIS-C ICD-10 code) are in Figure 4. The MIS-C group demonstrated a slight male predominance (59%) with a significant proportion of Hispanic and Black/African American children (25% and 33%, respectively). Of children with MIS-C, 12% required mechanical ventilation and 31% received vasopressors or inotropes.

**Figure 4:**
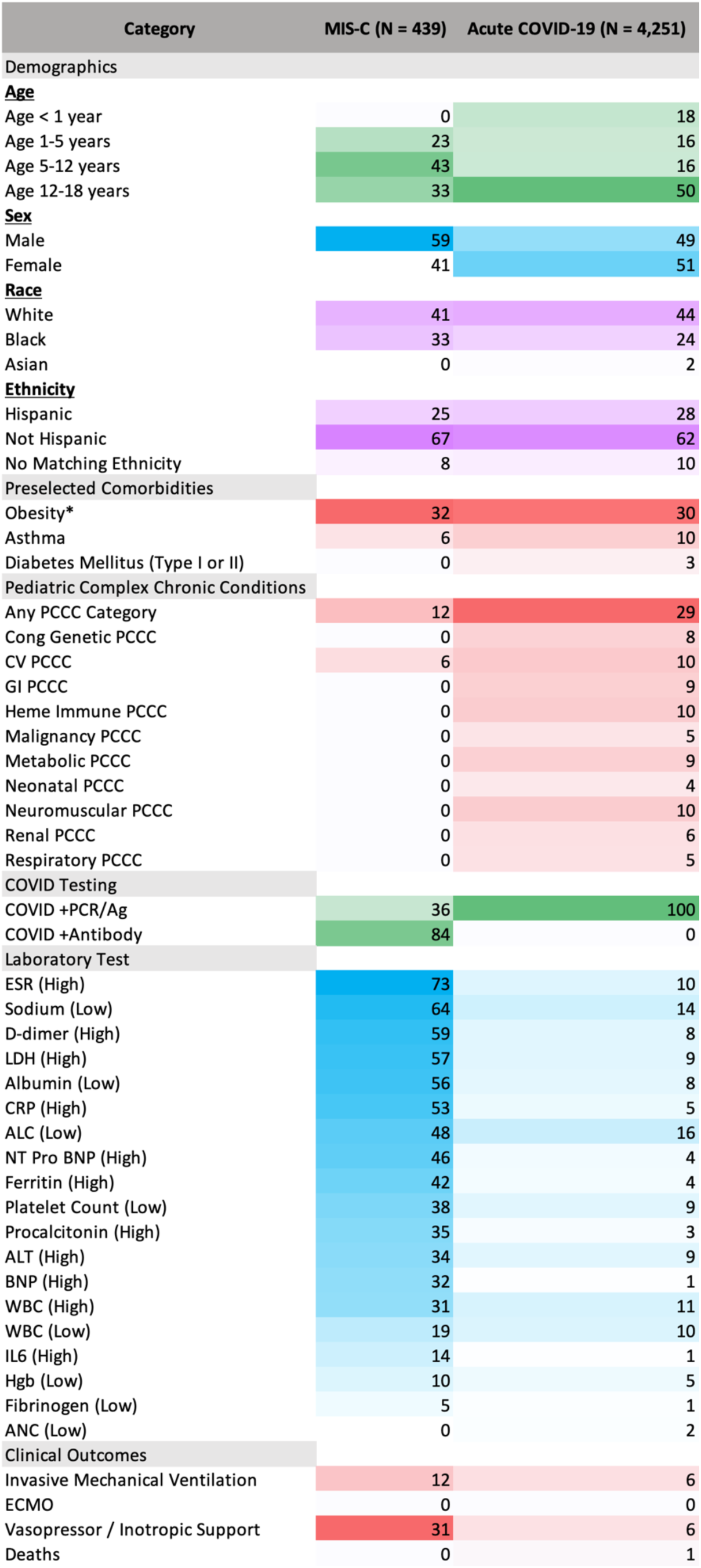
Characteristics and outcomes of children with MIS-C versus acute COVID-19. Heatmap comparing the percent of children in the MIS-C and acute COVID-19 subgroups with a given demographic characteristic, pre-existing comorbidity, abnormal lab value during hospitalization, or clinical outcome. See Supplemental Table 6 for the absolute number of patients in each corresponding category. *The percent of children with obesity was calculated by dividing the number of children ≥2-years-old who had a BMI for age and sex that was ≥95th percentile by the number of children in that subgroup who were ≥2 years old who had a BMI measurement available.

Compared to hospitalized children with acute COVID-19 only, hospitalized children with MIS-C were more likely to be 1-to-12-years-old, male, Black/African American, and less likely to have asthma, diabetes, or a PCCC (p < 0.04 for all). MIS-C patients were more likely to require vasoactive-inotropic support or invasive ventilation, and more likely to have a severe maximum clinical severity (p < 0.03)(Figure 4). Lastly, for 18 of the 19 preselected laboratory tests, the MIS-C group had a higher proportion of children with abnormal values compared to the acute COVID group (p < 0.001 for each).

## Discussion

In the largest U.S. SARS-CoV-2 positive pediatric cohort to date, we fill a gap in the pediatric SARS-CoV-2 literature: a description of vital sign and laboratory values and trajectories during hospitalization among infected children with varying peak clinical severity. Although reports from Feldstein et al.^5,6^ include median lab results from children with MIS-C and acute COVID-19, ours is the first report describing vital sign and laboratory trajectories during pediatric hospitalization with specific attention to maximum clinical severity. We observed notable differences in initial vital sign and laboratory test values between severity subgroups, which, combined with the observed intergroup differences in demographics and pre-existing comorbidities, suggest that early identification of children likely to progress to a more severe phenotype could be achieved using readily available data elements from the day of admission. Future work should include design of predictive models and clinical decision support tools geared toward early identification of children at high risk for subsequent deterioration and severe disease.

In addition, we report on changes in case and hospitalization rates, age and severity distributions, and treatment regimens throughout the study period. This cohort’s 5.7% hospitalization rate among positive cases is similar to the 7% rate in Bailey^14^ et al. although lower than that noted in Kompaniyets et al.^10^ (9.9%) and Preston et al.^4^ (12%). Our cohort’s median age was similar to that of Kompaniyets et al^10^ (12.5 years in our study vs 12 years) as was the prevalence of comorbid asthma (8% vs 10.2%) and neurologic/neuromuscular disease (3% vs 3.9%). Our cohort’s rate of mechanical ventilation (7%) among hospitalized patients was similar to two prior reports (7% and 6%)^7,10^ though less than that of Belay et al.^13^ (14.6% in the acute COVID-19 cohort). Similar to Gotzinger^7^ and Preston^4^ et al., we observed that male sex and the presence of a pre-existing chronic medical condition were risk factors for more severe disease. In addition, we found that Black/African American race and numerous other PCCC categories were similarly associated with higher maximum clinical severity once hospitalized.

Although Shekerdemian^9^ and Feldstein et al.^6^ both reported use of various antimicrobial and immunomodulatory medications in pediatric SARS-CoV-2, ours is the first report outlining differences in medication use over time. In this study, systemic antimicrobials and immunomodulatory medications were frequently used (especially in the severe subgroup) with antibacterials and systemic corticosteroids most commonly employed. The high frequency of steroid use, specifically, is likely related to emerging evidence of efficacy for treatment of severe COVID-19 and MIS-C^26,27^.

As others have reported^6,13^, compared to children with acute COVID-19, we found that children with MIS-C were more likely to be male, Black/African-American, and less likely to have pre-existing comorbidities (asthma, diabetes, or a PCCC). In our study, MIS-C cases also demonstrated a more severe clinical phenotype with significant laboratory evidence of inflammation and frequent need for invasive ventilation and vasoactive-inotropic support.

Given the recent introduction of ICD-10 codes for MIS-C (and unknown consistency of use), their sensitivity for MIS-C is uncertain. However, in the setting of a positive SARS-CoV-2 test, the positive predictive value is likely to be high. The MIS-C incidence we describe is likely a conservative estimate. Further work is needed to develop and validate computable phenotypes for identification of MIS-C cases within N3C and other large databases for subsequent research.

Clinical outcomes (e.g. rates of mechanical ventilation) and medication utilization in acute COVID-19 and MIS-C have varied between studies^6,7,9,10,14,28^. Many reports of pediatric acute COVID-19 and MIS-C originate from single centers or health systems ^2,3,28-31^. Recently, several large-scale studies reported risk factors and outcomes for pediatric COVID-19^4,7,9,10,14^ and MIS-C^6,13^. However, none have included analysis of highly granular, patient-level data to compare trajectories of individual vital sign and laboratory values between different clinical severity subgroups. Such analyses improve the capacity to build SARS-CoV-2 specific predictive models and clinical decision support tools.

This study has important limitations to consider. As data are aggregated from many health systems using four different CDMs that vary in granularity, some sites may have systematic missingness of certain variables. Additionally, some respiratory data (oxygen flow, FiO2, and specific ventilator settings) are not fully available. The exact timing of laboratory specimens is inconsistently provided by sites. As such, labs were standardized to a calendar day. Furthermore, with high rates of asymptomatic-to-minimally-symptomatic pediatrics infections and increasing adoption of universal SARS-CoV-2 testing policies for pediatric hospital admissions, we cannot definitively attribute reasons for hospital admissions (SARS-CoV-2 versus another unrelated cause). This may limit interpretation of variables associated with higher clinical severity. Additionally, given the low number of severe pediatric cases, interpretation of incidence over time is challenging. Lastly, although ICD-10 coding for MIS-C has likely improved with time and increased awareness of the condition, many cases of MIS-C (especially in the earlier months of the study period) were likely not identified as such, limiting interpretation of MIS-C subgroup analysis.

In summary, this study reports the characteristics and outcomes of the largest U.S. cohort of children with SARS-CoV-2 infection to date. The N3C database provides a geographically and demographically diverse, granular view of pediatric SARS-CoV-2 infections and allows for novel vital sign and laboratory value trajectory mapping. Further work is needed to optimize translation of this knowledge into improved clinical care.

## Data Availability

The N3C Data Enclave (covid.cd2h.org/enclave) houses fully reproducible, transparent, and broadly available limited and de-identified datasets (HIPAA definitions available at https://www.hhs.gov/hipaa/index.html). Enclave data can be accessed by investigators at institutions that have signed a Data Use Agreement with the NIH who have taken the required human subjects and security training and agree to the N3C User Code of Conduct. Those desiring access to the limited dataset need also supply an IRB protocol from their institution. Data access requests are reviewed by the NIH Data Access Committee. A full description of the N3C Enclave governance has been published (https://academic.oup.com/jamia/article/28/3/427/5893482) and additional information on the access application process is available on the NCATS website: https://ncats.nih.gov/n3c/about/applying-for-access. The data model utilized is OMOP 5.3.1, and specifications are available at: https://ncats.nih.gov/files/OMOP_CDM_COVID.pdf
Additional N3C resources including code and governance documents are available within the project GitHub repositories and/or in Zenodo for archival purposes: https://github.com/National-COVID-Cohort-Collaborative
https://zenodo.org/communities/cd2h-covid/

https://unite.nih.gov/

https://covid.cd2h.org/enclave

https://academic.oup.com/jamia/article/28/3/427/5893482

https://ncats.nih.gov/n3c/about/applying-for-access

https://ncats.nih.gov/files/OMOP_CDM_COVID.pdf

https://github.com/National-COVID-Cohort-Collaborative

https://zenodo.org/communities/cd2h-covid/

## Ethics and Regulatory

The N3C data transfer to NCATS is performed under a Johns Hopkins University Reliance Protocol # IRB00249128 or individual site agreements with NIH.

Use of the N3C data for this study is authorized under the following IRB Protocols:

| **Site** | **IRB name** | **Exempted vs approved** | **Protocol number** |
| --- | --- | --- | --- |
| University of Colorado | Colorado Multiple Institutional Review Board | approved | 20-2225 |
| Johns Hopkins University | Johns Hopkins Office of Human Subjects Research<br>- Institutional Review Board | approved | IRB00249128 |
| University of North Carolina | University of North Carolina Chapel Hill Institutional Review Board | exempted | 20-3106 |
| Stony Brook University | Office of Research Compliance, Division of Human Subject Protections, Stony Brook University | exempted | IRB2020-00604 |

## Contributions

Authorship was determined using ICMJE recommendations.

Concept and Design: Martin, Bennett, Moffitt, DeWitt, Russell, Pfaff, Saltz, Chute, Haendel Acquisition, Analysis, or Interpretation of Data: Martin, Bennett, Moffitt, DeWitt, Russell, Anand, Bradwell, Bremer, Gabriel, Girvin, Hajagos, McMurry, Wooldridge, Yoo, Gersing, Pfaff, Chute, Haendel

Drafting of the Manuscript: Martin, Bennett, Moffitt, DeWitt, Russell

Critical Revision of the Manuscript for Important Intellectual Content: Martin, Bennett, Moffitt, DeWitt, Russell, Anand, Bradwell, Bremer, Gabriel, Girvin, Hajagos, McMurry, Wooldridge, Yoo, Gersing, Pfaff, Chute, Haendel

Statistical Analysis: Martin, Bennett, Moffitt, DeWitt, Russell

Obtained Funding: McMurry, Walden, Gersing, Haendel, Chute, Bennett

Administrative, technical, or material support: Neumann, Walden, McMurry

Supervision: Bennett, Moffitt, Haendel, Chute

