## Supplemental Materials for "Children with SARS-CoV-2 in the National COVID Cohort Collaborative (N3C)"

### **Supplemental Methods**

*N3C Structure*

As described in prior work^1-3^, the N3C is housed in a cloud-based, secure, data enclave managed by the National Center for Advancing Translational Sciences (NCATS) to which contributing partner sites submit a limited data set in one of four common data models (CDMs). Thereafter, sites continue to provide weekly data updates. N3C harmonizes all data into the Observational Medical Outcomes Partnership (OMOP) CDM^4^. See Haendel et al.^3^ for details regarding data transfer, harmonization, and data quality assurance and integration. Computable phenotypes for COVID-19 and N3C COVID-negative controls are publicly available on GitHub^1^.

*Included Patients and Prior Medical History Data*

N3C includes cases and controls available for analysis including outpatient and inpatient visits. N3C includes encounters after 1/1/2020 from study sites who meet specific SARS-CoV-2 testing and diagnosis code criteria. The database includes any patient from an N3C study site with an encounter after 1/1/2020 that includes:

1) one of a set of a priori-defined SARS-CoV-2 laboratory tests,
2) a “strong positive” diagnostic code, or
3) two “weak positive” diagnostic codes during one encounter or on the same date prior to 5/1/2020.

As noted above, the cohort definition is publicly available on GitHub^1^. Encounters in the same health system on or after 1/1/2018 are also included to provide relevant clinical information on pre-existing health conditions (“lookback data”).

*Index Encounter and Clinical Severity Definition*

The N3C index encounter assigned to a child with positive SARS-CoV-2 testing was identified via evaluation of all encounters starting up to 30 days before through 7 days after each positive test result. We selected as the index encounter that which, amongst this encounter set, demonstrated the maximum Clinical Progression Scale (CPS) score (created by the World Health Organization [WHO] for COVID-19 clinical research^5^). Of note, we combined certain WHO CPS categories due to data limitations (e.g., certain sites did not submit fraction of inspired oxygen [FiO_2_]), resulting in four separate outcome groups:

1) Mild (outpatient visit only)
2) Mild ED (outpatient with emergency department visit)
3) Moderate (hospitalized without requiring invasive ventilation, vasopressors/inotropes, or ECMO)
4) Severe (hospitalized and requiring invasive ventilation, vasopressors/inotropes, or ECMO, death, or discharge to hospice)

As such, we identified a single encounter for each SARS-CoV-2 positive pediatric patient for cohort analysis. All subsequent visit-level analyses were performed using this select encounter only.

*Comorbidity Identification*

All concept sets developed for and utilized within this manuscript are freely available on the platform and include input from clinical subject matter and informatics experts. Children with asthma, obesity, and diabetes mellitus as well as those who received vasoactive-inotropic support were identified via adaptation and application of existing concept sets within the enclave. Children with a pediatric complex chronic condition^6^ (PCCC) were identified using our prior PCCC R implementation^7^.

*Additional Software Packages Used*

We used a variety of SQL, python, and R packages to clean and analyze data. We utilized SparkR^8^ and pyspark^9^ to interface with Apache Spark^10^ and utilized R’s ggplot2^11^ for all visualizations.

*Declaration of Interests / Conflict of Interest Disclosures*

Dr. Bennett reported receiving grants from the National Institutes of Health (NIH)/National Center for Advancing Translational Sciences (NCATS) during the conduct of the study and grants from the NIH/Eunice Kennedy Shriver National Institute of Child Health and Human Development and NIH/National Institute of Allergy and Infectious Diseases outside the submitted work. Dr. Moffitt reported receiving grants from the NIH during the conduct of the study. Dr. Hajagos reported receiving grants from the NIH/NCATS during the conduct of the study. Dr. Bradwell reported being employed by Palantir Technologies during the conduct of the study and outside the submitted work. Ms. Gabriel reported receiving grants from the NIH/NCATS during the conduct of the study. Dr. Girvin reported being an employee of Palantir Technologies. Ms. McMurry reported being a cofounder of Pryzm Health outside the submitted work. Dr. Pfaff reported receiving grants from the NIH/NCATS during the conduct of the study. Dr. Haendel reported receiving grants from the NIH during the conduct of the study. Dr. Chute reported receiving grants from the NIH/NCATS during the conduct of the study. No conflicts of interest reported for all other authors.

*Funding / Support:*

As described in Bennett et al.^2^, the primary study sponsors are multiple institutes of the U.S. National Institutes of Health. The National Center for Advancing Translational Sciences is the primary steward of the N3C data and created the underlying architecture of the N3C Data Enclave, manages the Data Transfer Agreements and Data Use Agreements, houses the Data Access Committee, and supports contracts to vendors to help build various aspects of the N3C Data Enclave. Employees of the NIH and of the contracting companies are included as authors of the manuscript and participated in the writing and decision to submit the manuscript. Please see the Declaration of Interests / Conflict of Interest Disclosures section above.

The analyses for this work were conducted using data and tools accessed through the NCATS N3C Data Enclave and supported by NCATS U24 TR002306 (Drs Haendel, Chute, and Saltz; also supporting Dr. Pfaff, Ms. Walden, Ms. McMurry, Mr. Neumann, and Ms. Gabriel). This study was also supported by the following (institutions with release data) grants: U24TR002306 (Stony Brook University), U54GM104938 (Oklahoma Clinical and Translational Science Institute, University of Oklahoma Health Sciences Center), U54GM104942 (West Virginia Clinical and Translational Science Institute, West Virginia University), U54GM115428 (Mississippi Center for Clinical and Translational Research, University of Mississippi Medical Center), U54GM115458 (Great Plains IDeA-Clinical & Translational Research, University of Nebraska Medical Center), U54GM115516 (Northern New England Clinical & Translational Research Network, Maine Medical Center), UL1TR001420 (Wake Forest Clinical and Translational Science Institute, Wake Forest University Health Sciences), UL1TR001422 (Northwestern University Clinical and Translational Science Institute, Northwestern University), UL1TR001425 (Center for Clinical and Translational Science and Training, University of Cincinnati), UL1TR001439 (Institute for Translational Sciences, University of Texas Medical Branch at Galveston), UL1TR001450 (South Carolina Clinical & Translational Research Institute, Medical University of South Carolina), UL1TR001453 (UMass Center for Clinical and Translational Science, University of Massachusetts Medical School Worcester), UL1TR001855 (Southern California Clinical and Translational Science Institute, University of Southern California), UL1TR001873 (Irving Institute for Clinical and Translational Research, Columbia University Irving Medical Center), UL1TR001876 (Clinical and Translational Science Institute at Children’s National, George Washington Children’s Research Institute), UL1TR001998 (Appalachian Translational Research Network, University of Kentucky), UL1TR002001 (University of Rochester Clinical & Translational Science Institute), UL1TR002003 (University of Illinois at Chicago Center for Clinical and Translational Science), UL1TR002014 (Penn State Clinical and Translational Science Institute), UL1TR002240 (Michigan Institute for Clinical and Health Research, University of Michigan at Ann Arbor), UL1TR002243 (Vanderbilt Institute for Clinical and Translational Research, Vanderbilt University Medical Center), UL1TR002319 (Institute of Translational Health Sciences, University of Washington), UL1TR002345 (Institute of Clinical and Translational Sciences, Washington University in St. Louis), UL1TR002369 (Oregon Clinical and Translational Research Institute, Oregon Health & Science University), UL1TR002373 (Wisconsin Network for Health Research, University of Wisconsin-Madison), UL1TR002389 (Institute for Translational Medicine, Rush University Medical Center), UL1TR002389 (Institute for Translational Medicine, University of Chicago), UL1TR002489 (North Carolina Translational and Clinical Science Institute, University of North Carolina at Chapel Hill), UL1TR002494 (Clinical and Translational Science Institute, University of Minnesota), UL1TR002535 (Colorado Clinical and Translational Sciences Institute and Children’s Hospital Colorado), UL1TR002537 and UL1TR002535 03S2 (Institute for Clinical and Translational Science, University of Iowa), UL1TR002538 (Uhealth Center for Clinical and Translational Science, University of Utah), UL1TR002544 (Tufts Clinical and Translational Science Institute, Tufts Medical Center), UL1TR002553 (Duke Clinical and Translational Science Institute, Duke University), UL1TR002649 (C. Kenneth and Dianne Wright Center for Clinical and Translational Research, Virginia Commonwealth University), UL1TR002733 (Center for Clinical and Translational Science, Ohio State University), UL1TR002736 (University of Miami Clinical and Translational Science Institute), UL1TR003015 (iTHRIVL Integrated Translational health Research Institute of Virginia, University of Virginia), UL1TR003015 (iTHRIVL Integrated Translational health Research Institute of Virginia, Carilion Clinic), UL1TR003096 (Center for Clinical and Translational Science, University of Alabama at Birmingham), UL1TR003098 (Johns Hopkins Institute for Clinical and Translational Research, Johns Hopkins University), UL1TR003107 (Consortium of Rural States, University of Arkansas for Medical Sciences), U54GM104941 (Delaware CTR ACCEL Program, Nemours), UL1TR002535 (Colorado Clinical and Translational Sciences Institute, University of Colorado, Denver, Anschutz Medical Campus), UL1TR002377 (Mayo Clinic Center for Clinical and Translational Science, Mayo Clinic Rochester), UL1TR002389 (Institute for Translational Medicine, Loyola University Medical Center), UL1TR002389 (Institute for Translational Medicine, Advocate Health Care Network). Additional support (data release pending) as provided as follows: UL1TR001866 (Center for Clinical and Translational Science, Rockefeller University), UL1TR002550 (Scripps Research Translational Institute, The Scripps Research Institute), UL1TR002645 (Institute for Integration of Medicine and Science, University of Texas Health Science Center at San Antonio), UL1TR003167 (Center for Clinical and Translational Sciences, The University of Texas Health Science Center at Houston), UL1TR002389 (Institute for Translational Medicine, NorthShore University HealthSystem), UL1TR001863 (Yale Center for Clinical Investigation, Yale New Haven Hospital), UL1TR002378 (Georgia Clinical and Translational Science Alliance, Emory University), UL1TR002384 (Weill Cornell Medicine Clinical and Translational Science Center, Weill Medical College of Cornell University), UL1TR002556 (Institute for Clinical and Translational Research at Einstein and Montefiore, Montefiore Medical Center), UL1TR001436 (Clinical and Translational Science Institute of Southeast Wisconsin, Medical College of Wisconsin), UL1TR001449 (University of New Mexico Clinical and Translational Science Center, University of New Mexico Health Sciences Center), UL1TR001876 (Clinical and Translational Science Institute at Children’s National, George Washington University), UL1TR003142 (Spectrum: The Stanford Center for Clinical and Translational Research and Education, Stanford University), UL1TR002529 (Indiana Clinical and Translational Science Institute, Regenstrief Institute), UL1TR001425 (Center for Clinical and Translational Science and Training, Cincinnati Children’s Hospital Medical Center), UL1TR001430 (Boston University Clinical and Translational Science Institute, Boston University Medical Campus), U54GM104940 (Louisiana Clinical and Translational Science Center, University Medical Center New Orleans), UL1TR001412 (Clinical and Translational Science Institute, The State University of New York at Buffalo), UL1TR002373 (Wisconsin Network For Health Research, Aurora Health Care), U54GM115677 (Advance Clinical Translational Research, Brown University), UL1TR003017 (New Jersey Alliance for Clinical and Translational Science, Rutgers, The State University of New Jersey), UL1TR002389 (Institute for Translational Medicine, Loyola University Chicago), UL1TR001445 (Langone Health’s Clinical and Translational Science Institute, New York University Grossman School of Medicine), UL1TR001878 (Institute for Translational Medicine and Therapeutics, Children’s Hospital of Philadelphia), UL1TR002366 (Frontiers: University of Kansas Clinical and Translational Science Institute, University of Kansas Medical Center), UL1TR002541 (Harvard Catalyst, Massachusetts General Brigham), INV-018455 (Bill and Melinda Gates Foundation grant to Sage Bionetworks, OCHIN), UL1TR001433 (ConduITS Institute for Translational Sciences, Icahn School of Medicine at Mount Sinai), U54GM104940 (Louisiana Clinical and Translational Science Center, Ochsner Medical Center), UL1TR001414 (University of California, Irvine Institute for Clinical and Translational Science), UL1TR001442 (Altman Clinical and Translational Research Institute, University of California, San Diego), UL1TR001860 (UCDavis Health Clinical and Translational Science Center, University of California, Davis), UL1TR001872 (University of California, San Francisco Clinical and Translational Science Institute), and UL1TR001881 (UCLA Clinical Translational Science Institute).

*Data Sharing*

The N3C Data Enclave (covid.cd2h.org/enclave) houses fully reproducible, transparent, and broadly available limited and de-identified datasets (HIPAA definitions available at https://www.hhs.gov/hipaa/index.html). Enclave data can be accessed by investigators at institutions that have signed a Data Use Agreement with the NIH who have taken the required human subjects and security training and agree to the N3C User Code of Conduct. Those desiring access to the limited dataset need also supply an IRB protocol from their institution. Data access requests are reviewed by the NIH Data Access Committee. A full description of the N3C Enclave governance has been published^3^ and additional information on the access application process is available on the NCATS website: https://ncats.nih.gov/n3c/about/applying-for-access. The data model utilized is OMOP 5.3.1, and specifications are available at: https://ncats.nih.gov/files/OMOP_CDM_COVID.pdf

Additional N3C resources including code and governance documents are available within the project GitHub repositories and/or in Zenodo for archival purposes: https://github.com/National-COVID-Cohort-Collaborative

https://zenodo.org/communities/cd2h-covid/

Additional information regarding each of the source Common Data Models is available at:

OHDSI: https://ohdsi.org/

PCORNet: https://pcornet.org/

ACT: https://www.dbmi.pitt.edu/node/53983

TriNetX: https://trinetx.com/

**Supplemental Tables**

| A. Pediatric Patients |  |  |  |  |  |  |  |
| --- | --- | --- | --- | --- | --- | --- | --- |
| **Demographic Category** | **All Children Tested** | **PCR/Ag Positive** | **PCR/Ag Negative** | **No PCR/Ag Test** | **Ab Positive** | **Ab Negative** | **No Ab Test** |
| N | 728,047 | 88,122 | 408,826 | 230,965 | 4,031 | 6,240 | 717,773 |
| Age |  |  |  |  |  |  |  |
| Age (y), mean (SD) | 9.6 (6.1) | 11.3 (5.9) | 9.2 (6.1) | 9.8 (6.0) | 11.3 (5.6) | 11.3 (5.7) | 9.6 (6.1) |
| Age (y), median (IQR) | 9.7 (3.9, 15.4) | 12.5 (6.4, 16.5) | 8.9 (3.4, 14.9) | 10.0 (4.2, 15.5) | 12.1 (6.7, 16.2) | 12.4 (6.4, 16.3) | 9.7 (3.9, 15.4) |
| Gender |  |  |  |  |  |  |  |
| Male | 373,311 (51.3%) | 44,077 (50.0%) | 211,492 (51.7%) | 117,683 (51.0%) | 2,078 (51.6%) | 3,236 (51.9%) | 367,996 (51.3%) |
| Female | 354,341 (48.7%) | 43,983 (49.9%) | 197,273 (48.3%) | 113,010 (48.9%) | 1,950 (48.4%) | 3,004 (48.1%) | 349,385 (48.7%) |
| Other | 395 (0.1%) | 62 (0.1%) | 61 (0.0%) | 272 (0.1%) | <20 (0%) | <20 (0%) | 392 (0.1%) |
| Ethnicity |  |  |  |  |  |  |  |
| Hispanic or Latino | 131,228 (18.0%) | 21,513 (24.4%) | 72,577 (17.8%) | 37,094 (16.1%) | 910 (22.6%) | 828 (13.3%) | 129,488 (18.0%) |
| Not Hispanic or  Latino | 497,126 (68.3%) | 54,481 (61.8%) | 293,512 (71.8%) | 149,053 (64.5%) | 2,768 (68.7%) | 4,942 (79.2%) | 489,415 (68.2%) |
| Missing/Unknown | 99,693 (13.7%) | 12,128 (13.8%) | 42,737 (10.5%) | 44,818 (19.4%) | 353 (8.8%) | 470 (7.5%) | 98,870 (13.8%) |
| Race |  |  |  |  |  |  |  |
| Asian | 17,543 (2.4%) | 1,989 (2.3%) | 10,430 (2.6%) | 5,121 (2.2%) | 92 (2.3%) | 222 (3.6%) | 17,229 (2.4%) |
| Black or African  American | 113,366 (15.6%) | 12,807 (14.5%) | 66,461 (16.3%) | 34,074 (14.8%) | 379 (9.4%) | 610 (9.8%) | 112,376 (15.7%) |
| Native Hawaiian /  Pacific Islander | 1,607 (0.2%) | 244 (0.3%) | 777 (0.2%) | 586 (0.3%) | <20 (0%) | <20 (0%) | 1,598 (0.2%) |
| White | 414,155 (56.9%) | 49,109 (55.7%) | 244,195 (59.7%) | 120,783 (52.3%) | 1,889 (46.9%) | 3,771 (60.4%) | 408,493 (56.9%) |
| Other | 168,594 (23.2%) | 22,343 (25.4%) | 81,847 (20.0%) | 64,365 (27.9%) | 1,637 (40.6%) | 1,580 (25.3%) | 165,377 (23.0%) |
| Missing/Unknown | 12,423 (1.7%) | 1,541 (1.7%) | 4,886 (1.2%) | 5,996 (2.6%) | 22 (0.5%) | 44 (0.7%) | 12,357 (1.7%) |
| B. Adult Patients |  |  |  |  |  |  |  |
| **Demographic Category** | **All Adults Tested** | **PCR/Ag Positive** | **PCR/Ag Negative** | **No PCR/Ag Test** | **Ab Positive** | **Ab Negative** | **No Ab Test** |
| N | 4,657,150 | 609,734 | 2,581,342 | 1,465,317 | 50,394 | 107,979 | 4,498,709 |
| Age |  |  |  |  |  |  |  |
| Age(y) mean (SD) | 49.9 (18.5) | 48.1 (18.6) | 50.7 (18.6) | 49.5 (18.3) | 49.9 (16.9) | 49.7 (16.4) | 50.0 (18.6) |
| Age(y) median (IQR) | 49.7 (34.1, 64.0) | 47.0 (32.0, 61.6) | 50.6 (34.6, 64.9) | 49.0 (34.1, 63.1) | 50.1 (36.1, 62.2) | 49.3 (36.2, 61.9) | 49.7 (34.0, 64.0) |
| Gender |  |  |  |  |  |  |  |
| Male | 2,017,882 (43.3%) | 275,381 (45.2%) | 1,099,725 (42.6%) | 642,454 (43.8%) | 20,254 (40.2%) | 42,888 (39.7%) | 1,954,708 (43.5%) |
| Female | 2,635,475 (56.6%) | 333,944 (54.8%) | 1,480,364 (57.3%) | 820,733 (56.0%) | 30,022 (59.6%) | 65,071 (60.3%) | 2,540,346 (56.5%) |
| Other | 3,793 (0.1%) | 409 (0.1%) | 1,253 (0.0%) | 2,130 (0.1%) | 118 (0.2%) | 20 (0.0%) | 3,655 (0.1%) |
| Ethnicity |  |  |  |  |  |  |  |
| Hispanic or Latino | 488,543 (10.5%) | 103,346 (16.9%) | 250,517 (9.7%) | 134,521 (9.2%) | 6,469 (12.8%) | 8,056 (7.5%) | 474,017 (10.5%) |
| Not Hispanic or  Latino | 3,598,581 (77.3%) | 433,373 (71.1%) | 2,104,378 (81.5%) | 1,060,293 (72.4%) | 38,475 (76.3%) | 91,119 (84.4%) | 3,468,949 (77.1%) |
| Missing/Unknown | 570,026 (12.2%) | 73,015 (12.0%) | 226,447 (8.8%) | 270,503 (18.5%) | 5,450 (10.8%) | 8,804 (8.2%) | 555,743 (12.4%) |
| Race |  |  |  |  |  |  |  |
| Asian | 138,094 (3.0%) | 17,974 (2.9%) | 81,898 (3.2%) | 38,185 (2.6%) | 2,078 (4.1%) | 4,950 (4.6%) | 131,066 (2.9%) |
| Black or African  American | 695,570 (14.9%) | 101,450 (16.6%) | 423,086 (16.4%) | 170,925 (11.7%) | 5,598 (11.1%) | 9,680 (9.0%) | 680,287 (15.1%) |
| Native Hawaiian /  Pacific Islander | 7,756 (0.2%) | 1,251 (0.2%) | 3,895 (0.2%) | 2,609 (0.2%) | 62 (0.1%) | 154 (0.1%) | 7,540 (0.2%) |
| White | 2,940,899 (63.1%) | 360,319 (59.1%) | 1,706,871 (66.1%) | 873,299 (59.6%) | 29,512 (58.6%) | 75,678 (70.1%) | 2,835,672 (63.0%) |
| Other | 838,887 (18.0%) | 123,799 (20.3%) | 355,243 (13.8%) | 359,647 (24.5%) | 12,974 (25.7%) | 17,119 (15.9%) | 808,768 (18.0%) |
| Missing/Unknown | 33,122 (0.7%) | 4,155 (0.7%) | 8,635 (0.3%) | 20,330 (1.4%) | 97 (0.2%) | 301 (0.3%) | 32,724 (0.7%) |

Supplemental Table 1: N3C Cohort Characteristics. Demographic characteristics for pediatric (A) and adult (B) N3C patients stratified SARS-CoV-2 test type and result. Number of patients with “unknown result” for PCR/Ag or Ab testing not shown. Per N3C policy, we censored any cells with <20 patients and replaced each with “<20 (0%).”

*Definitions:* Ab = antibody, Ag = antigen, IQR interquartile range, PCR = polymerase chain reaction, SD = standard deviation

| **Medication Category** | **Medication Name** | **Severity Category** | **Children who received the medication, n (%)** | **p value*** |
| --- | --- | --- | --- | --- |
| Antimicrobial Overall |  | Moderate | 1447 (32) | p < 0.001 |
|  |  | Severe | 484 (71) |  |
| Immunomodulation  Overall |  | Moderate | 702 (16) | p < 0.001 |
|  |  | Severe | 365 (53) |  |
| Antimicrobial | Azithromycin | Moderate | 147 (3) | p < 0.001 |
|  |  | Severe | 39 (6) |  |
| Antimicrobial | Remdesivir | Moderate | 86 (2) | p < 0.001 |
|  |  | Severe | 62 (9) |  |
| Antimicrobial | Systemic  Antibacterial | Moderate | 1386 (31) | p < 0.001 |
|  |  | Severe | 466 (68) |  |
| Antimicrobial | Systemic Antifungal | Moderate | 60 (1) | p < 0.001 |
|  |  | Severe | 76 (11) |  |
| Antimicrobial | Systemic Antiviral | Moderate | 142 (3) | p < 0.001 |
|  |  | Severe | 99 (14) |  |
| Immunomodulation | Anakinra | Moderate | 34 (1) | p < 0.001 |
|  |  | Severe | 65 (9) |  |
| Immunomodulation | Dexamethasone | Moderate | 192 (4) | p < 0.001 |
|  |  | Severe | 88 (13) |  |
| Immunomodulation | Hydrocortisone | Moderate | 93 (2) | p < 0.001 |
|  |  | Severe | 136 (20) |  |
| Immunomodulation | Infliximab | Moderate | 28 (1) | *N/A* |
|  |  | Severe | <20 (0) |  |
| Immunomodulation | Methylprednisolone | Moderate | 239 (5) | p < 0.001 |
|  |  | Severe | 137 (20) |  |
| Immunomodulation | Prednisone | Moderate | 296 (7) | p < 0.001 |
|  |  | Severe | 139 (20) |  |
| Immunomodulation | Systemic  Corticosteroid | Moderate | 661 (15) | p < 0.001 |
|  |  | Severe | 343 (50) |  |

Supplemental Table 2: Statistical comparison of medication administration during pediatric hospitalization among moderate (N = 4,528 ) and severe (N = 685) subgroups.

*Comparison of the proportions of patients in the moderate vs severe subgroups who received a given medication / medication class.

| **Vital Sign** | **Severity Comparison Metric** | **Mean Value** | **Standard Error** | **CI Lower Limit** | **CI Upper Limit** | **p value** |
| --- | --- | --- | --- | --- | --- | --- |
| Systolic blood pressure  (mmHg) | Severe: Day 0 | 103.2 | 0.8 | 101.6 | 104.8 |  |
|  | Severe: Day 7 | 109.4 | 1.2 | 107.1 | 111.7 |  |
|  | Severe: Day 7 - Day 0 | 6.2 | 1.4 | 3.4 | 9.0 | p < 0.001 |
|  | Moderate: Day 0 | 108.4 | 0.3 | 107.8 | 109.1 |  |
|  | Moderate: Day 7 | 112.0 | 0.8 | 110.3 | 113.6 |  |
|  | Moderate: Day 7 - Day 0 | 3.5 | 1.0 | 1.6 | 5.5 | p < 0.001 |
|  | Severe Day 0 - Moderate Day 0 | -6.2 | 0.88 | -7.92 | -4.47 | p < 0.001 |
| Heart rate  (beats per minute) | Severe: Day 0 | 115.2 | 1.3 | 112.7 | 117.7 |  |
|  | Severe: Day 7 | 92.1 | 2.0 | 88.3 | 96.0 |  |
|  | Severe: Day 7 - Day 0 | -23.1 | 2.2 | -27.4 | -18.7 | p < 0.001 |
|  | Moderate: Day 0 | 112.8 | 0.6 | 111.6 | 114.1 |  |
|  | Moderate: Day 7 | 85.9 | 1.5 | 82.9 | 88.9 |  |
|  | Moderate: Day 7 - Day 0 | -26.9 | 1.7 | -30.2 | -23.5 | p < 0.001 |
|  | Severe Day 0 - Moderate Day 0 | 4.69 | 1.52 | 1.72 | 7.66 | p = 0.002 |
| Respiratory rate  (breaths per minute) | Severe: Day 0 | 27.1 | 0.5 | 26.1 | 28.2 |  |
|  | Severe: Day 7 | 24.2 | 0.8 | 22.7 | 25.8 |  |
|  | Severe: Day 7 - Day 0 | -2.9 | 1.1 | -5.0 | -0.8 | p = 0.006 |
|  | Moderate: Day 0 | 25.3 | 0.2 | 24.9 | 25.7 |  |
|  | Moderate: Day 7 | 21.9 | 0.4 | 21.0 | 22.8 |  |
|  | Moderate: Day 7 - Day 0 | -3.4 | 0.5 | -4.4 | -2.4 | p < 0.001 |
|  | Severe Day 0 - Moderate Day 0 | 2.05 | 0.5 | 1.06 | 3.04 | p < 0.001 |
| Diastolic blood pressure  (mmHg) | Severe: Day 0 | 59.9 | 0.6 | 58.7 | 61.0 |  |
|  | Severe: Day 7 | 62.9 | 0.9 | 61.0 | 64.7 |  |
|  | Severe: Day 7 - Day 0 | 3.0 | 1.2 | 0.6 | 5.4 | p = 0.01 |
|  | Moderate: Day 0 | 64.7 | 0.2 | 64.2 | 65.2 |  |
|  | Moderate: Day 7 | 67.9 | 0.7 | 66.6 | 69.1 |  |
|  | Moderate: Day 7 - Day 0 | 3.2 | 0.8 | 1.6 | 4.7 | p < 0.001 |
|  | Severe Day 0 - Moderate Day 0 | -5.1 | 0.65 | -6.37 | -3.83 | p < 0.001 |
| Mean arterial pressure  (mmHg) | Severe: Day 0 | 70.4 | 1.4 | 67.7 | 73.1 |  |
|  | Severe: Day 7 | 74.3 | 1.7 | 70.9 | 77.7 |  |
|  | Severe: Day 7 - Day 0 | 3.9 | 1.9 | 0.2 | 7.7 | p = 0.04 |
|  | Moderate: Day 0 | 74.3 | 0.7 | 72.9 | 75.8 |  |
|  | Moderate: Day 7 | 82.8 | 1.6 | 79.6 | 86.0 |  |
|  | Moderate: Day 7 - Day 0 | 8.4 | 2.0 | 4.5 | 12.4 | p < 0.001 |
|  | Severe Day 0 - Moderate Day 0 | -7.6 | 1.99 | -11.5 | -3.69 | p < 0.001 |
| Temperature  (℃) | Severe: Day 0 | 37.2 | 0.0 | 37.1 | 37.3 |  |
|  | Severe: Day 7 | 36.7 | 0.1 | 36.6 | 36.8 |  |
|  | Severe: Day 7 - Day 0 | -0.5 | 0.1 | -0.7 | -0.3 | p < 0.001 |
|  | Moderate: Day 0 | 37.2 | 0.0 | 37.1 | 37.2 |  |
|  | Moderate: Day 7 | 36.6 | 0.0 | 36.5 | 36.6 |  |
|  | Moderate: Day 7 - Day 0 | -0.6 | 0.0 | -0.7 | -0.5 | p < 0.001 |
|  | Severe Day 0 - Moderate Day 0 | -0.03 | 0.05 | -0.13 | 0.07 | p = 0.55 |
| SpO2  (%) | Severe: Day 0 | 95.6 | 0.3 | 94.9 | 96.3 |  |
|  | Severe: Day 7 | 95.1 | 0.5 | 94.1 | 96.0 |  |
|  | Severe: Day 7 - Day 0 | -0.6 | 0.6 | -1.8 | 0.7 | p = 0.37 |
|  | Moderate: Day 0 | 97.7 | 0.1 | 97.5 | 97.9 |  |
|  | Moderate: Day 7 | 97.6 | 0.2 | 97.2 | 97.9 |  |
|  | Moderate: Day 7 - Day 0 | -0.1 | 0.2 | -0.5 | 0.3 | p = 0.50 |
|  | Severe Day 0 - Moderate Day 0 | -2.63 | 0.43 | -3.48 | -1.78 | p < 0.001 |

Supplemental Table 3: Statistical comparison of the initial values and trends (from day 0 to day 7 of hospitalization) for vital signs between moderate and severe pediatric subgroups.

*Abbreviations:* CI = confidence interval, SpO2 = peripheral oxygen saturation

| **Laboratory Test** | **Severity Comparison Metric** | **Mean** | **Standard Error** | **CI Lower** | **CI Upper** | **p value** |
| --- | --- | --- | --- | --- | --- | --- |
| Albumin (g/dL) | Severe Day 7 - Day 0 | -0.2 | 0.0 | -0.3 | -0.1 | p < 0.001 |
|  | Moderate Day 7 - Day 0 | -0.6 | 0.0 | -0.6 | -0.5 | p < 0.001 |
|  | Severe Day 0 - Moderate Day 0 | -0.6 | 0.0 | -0.7 | -0.5 | p < 0.001 |
| ALT (SGPT), IU/L | Severe Day 7 - Day 0 | -132.4 | 110.1 | -348.3 | 83.4 | p = 0.23 |
|  | Moderate Day 7 - Day 0 | -103.1 | 45.4 | -192.1 | -14.2 | p = 0.02 |
|  | Severe Day 0 - Moderate Day 0 | 25.1 | 9.4 | 6.7 | 43.5 | p = 0.008 |
| AST (SGOT), IU/L | Severe Day 7 - Day 0 | -72.8 | 45.5 | -162.0 | 16.4 | p = 0.11 |
|  | Moderate Day 7 - Day 0 | -22.1 | 9.2 | -40.2 | -3.9 | p = 0.02 |
|  | Severe Day 0 - Moderate Day 0 | 31.2 | 8.0 | 15.5 | 46.8 | p < 0.001 |
| Bilirubin  (Unconjugated / Indirect), mg/dL | Severe Day 7 - Day 0 | -0.2 | 0.1 | -0.5 | 0.0 | p = 0.02 |
|  | Moderate Day 7 - Day 0 | -0.3 | 0.1 | -0.5 | 0.0 | p = 0.03 |
|  | Severe Day 0 - Moderate Day 0 | -0.1 | 0.2 | -0.3 | 0.2 | p = 0.70 |
| Bilirubin  (Conjugated / Direct),mg/dL | Severe Day 7 - Day 0 | -0.1 | 0.3 | -0.7 | 0.5 | p = 0.84 |
|  | Moderate Day 7 - Day 0 | -1.6 | 3.1 | -7.6 | 4.5 | p = 0.61 |
|  | Severe Day 0 - Moderate Day 0 | 0.2 | 0.1 | -0.1 | 0.4 | p = 0.12 |
| Bilirubin (total), mg/dL | Severe Day 7 - Day 0 | -0.3 | 0.3 | -0.8 | 0.2 | p = 0.25 |
|  | Moderate Day 7 - Day 0 | -0.9 | 0.2 | -1.3 | -0.4 | p < 0.001 |
|  | Severe Day 0 - Moderate Day 0 | 0.2 | 0.1 | 0.0 | 0.5 | p = 0.10 |
| BNP (all forms), pg/mL | Severe Day 7 - Day 0 | -6273.3 | 1265.8 | -8754.1 | -3792.5 | p < 0.001 |
|  | Moderate Day 7 - Day 0 | -1208.3 | 476.8 | -2142.7 | -273.8 | p = 0.01 |
|  | Severe Day 0 - Moderate Day 0 | 5669.8 | 1106.8 | 3500.5 | 7839.0 | p < 0.001 |
| BUN, mg/dL | Severe Day 7 - Day 0 | -0.3 | 2.1 | -4.4 | 3.7 | p = 0.87 |
|  | Moderate Day 7 - Day 0 | -4.6 | 1.6 | -7.7 | -1.4 | p = 0.004 |
|  | Severe Day 0 - Moderate Day 0 | 5.3 | 0.7 | 3.8 | 6.7 | p < 0.001 |
| BUN/Creatinine ratio | Severe Day 7 - Day 0 | 12.9 | 2.6 | 7.9 | 18.0 | p < 0.001 |
|  | Moderate Day 7 - Day 0 | 6.2 | 2.0 | 2.2 | 10.1 | p = 0.002 |
|  | Severe Day 0 - Moderate Day 0 | 1.9 | 1.9 | -1.9 | 5.7 | p = 0.33 |
| c-reactive protein (CRP),  mg/L | Severe Day 7 - Day 0 | -145.2 | 11.9 | -168.6 | -121.7 | p < 0.001 |
|  | Moderate Day 7 - Day 0 | -99.6 | 8.3 | -115.9 | -83.4 | p < 0.001 |
|  | Severe Day 0 - Moderate Day 0 | 70.2 | 8.4 | 53.7 | 86.7 | p < 0.001 |
| Chloride, mmol/L | Severe Day 7 - Day 0 | -1.1 | 0.9 | -2.8 | 0.7 | p = 0.22 |
|  | Moderate Day 7 - Day 0 | 0.5 | 0.5 | -0.4 | 1.4 | p = 0.30 |
|  | Severe Day 0 - Moderate Day 0 | -0.3 | 0.3 | -0.9 | 0.3 | p = 0.32 |
| Creatinine, mg/dL | Severe Day 7 - Day 0 | -0.32 | 0.15 | -0.61 | -0.02 | p = 0.04 |
|  | Moderate Day 7 - Day 0 | -0.38 | 0.12 | -0.60 | -0.15 | p = 0.001 |
|  | Severe Day 0 - Moderate Day 0 | 0.26 | 0.07 | 0.13 | 0.39 | p < 0.001 |
| D-Dimer, mg/L FEU | Severe Day 7 - Day 0 | 0.6 | 0.8 | -0.9 | 2.1 | p = 0.46 |
|  | Moderate Day 7 - Day 0 | -1.0 | 0.5 | -2.1 | 0.0 | p = 0.05 |
|  | Severe Day 0 - Moderate Day 0 | 1.4 | 0.4 | 0.7 | 2.2 | p < 0.001 |
| Erythrocyte Sed. Rate, mm/hr | Severe Day 7 - Day 0 | 7.6 | 5.1 | -2.4 | 17.6 | p = 0.14 |
|  | Moderate Day 7 - Day 0 | 6.9 | 3.5 | 0.0 | 13.8 | p = 0.05 |
|  | Severe Day 0 - Moderate Day 0 | 6.5 | 2.5 | 1.5 | 11.5 | p = 0.01 |
| Ferritin, ng/mL | Severe Day 7 - Day 0 | 104.1 | 695.7 | -1259.5 | 1467.6 | p = 0.88 |
|  | Moderate Day 7 - Day 0 | 126.6 | 197.0 | -259.5 | 512.6 | p = 0.52 |
|  | Severe Day 0 - Moderate Day 0 | 375.6 | 140.5 | 100.3 | 650.9 | p = 0.008 |
| Fibrinogen | Severe Day 7 - Day 0 | -148.6 | 26.0 | -199.6 | -97.6 | p < 0.001 |
|  | Moderate Day 7 - Day 0 | -184.0 | 24.7 | -232.4 | -135.5 | p < 0.001 |
|  | Severe Day 0 - Moderate Day 0 | -33.3 | 18.4 | -69.2 | 2.7 | p = 0.07 |
| Glucose, mg/dL | Severe Day 7 - Day 0 | -15.1 | 4.6 | -24.2 | -6.0 | p = 0.001 |
|  | Moderate Day 7 - Day 0 | -35.1 | 6.4 | -47.6 | -22.6 | p < 0.001 |
|  | Severe Day 0 - Moderate Day 0 | 12.6 | 2.8 | 7.1 | 18.2 | p < 0.001 |
| Hemoglobin, g/dL | Severe Day 7 - Day 0 | -0.8 | 0.2 | -1.1 | -0.5 | p < 0.001 |
|  | Moderate Day 7 - Day 0 | -1.3 | 0.2 | -1.6 | -1.0 | p < 0.001 |
|  | Severe Day 0 - Moderate Day 0 | -0.7 | 0.1 | -0.9 | -0.4 | p < 0.001 |
| IL-6, pg/mL | Severe Day 7 - Day 0 | -94.0 | 74.5 | -240.1 | 52.0 | p = 0.21 |
|  | Moderate Day 7 - Day 0 | -97.0 | 27.7 | -151.4 | -42.7 | p < 0.001 |
|  | Severe Day 0 - Moderate Day 0 | 86.1 | 37.7 | 12.3 | 159.9 | p = 0.02 |
| Lactate Dehydrogenase  (LDH), units/L | Severe Day 7 - Day 0 | -267.0 | 124.2 | -510.5 | -23.5 | p = 0.03 |
|  | Moderate Day 7 - Day 0 | -141.8 | 54.3 | -248.3 | -35.3 | p = 0.009 |
|  | Severe Day 0 - Moderate Day 0 | 172.0 | 58.3 | 57.7 | 286.2 | p = 0.003 |
| Lactate, mM | Severe Day 7 - Day 0 | -1.6 | 0.3 | -2.1 | -1.1 | p < 0.001 |
|  | Moderate Day 7 - Day 0 | -1.8 | 0.7 | -3.2 | -0.3 | p = 0.02 |
|  | Severe Day 0 - Moderate Day 0 | 0.6 | 0.1 | 0.4 | 0.9 | p < 0.001 |
| Lymphocytes (absolute),  x10^3^/$\mu$L | Severe Day 7 - Day 0 | 0.0 | 0.3 | -0.6 | 0.6 | p = 0.95 |
|  | Moderate Day 7 - Day 0 | 0.5 | 0.1 | 0.2 | 0.8 | p < 0.001 |
|  | Severe Day 0 - Moderate Day 0 | -0.5 | 0.1 | -0.7 | -0.3 | p < 0.001 |
| Lymphocytes (relative), % | Severe Day 7 - Day 0 | 4.5 | 1.1 | 2.4 | 6.6 | p < 0.001 |
|  | Moderate Day 7 - Day 0 | 10.1 | 1.0 | 8.1 | 12.0 | p < 0.001 |
|  | Severe Day 0 - Moderate Day 0 | -8.2 | 0.9 | -9.9 | -6.5 | p < 0.001 |
| Neutrophils (absolute),  x10^3^/$\mu$L | Severe Day 7 - Day 0 | 0.5 | 0.7 | -0.9 | 2.0 | p = 0.48 |
|  | Moderate Day 7 - Day 0 | -1.3 | 0.5 | -2.4 | -0.2 | p = 0.02 |
|  | Severe Day 0 - Moderate Day 0 | 2.1 | 0.4 | 1.3 | 2.8 | p < 0.001 |
| Neutrophils (relative), % | Severe Day 7 - Day 0 | -3.8 | 1.7 | -7.2 | -0.4 | p = 0.03 |
|  | Moderate Day 7 - Day 0 | -8.6 | 1.2 | -11.0 | -6.1 | p < 0.001 |
|  | Severe Day 0 - Moderate Day 0 | 6.5 | 1.4 | 3.8 | 9.2 | p < 0.001 |
| NT-proBNP, pg/mL | Severe Day 7 - Day 0 | -10951.6 | 2397.7 | -15651.0 | -6252.2 | p < 0.001 |
|  | Moderate Day 7 - Day 0 | -1690.1 | 652.9 | -2969.7 | -410.5 | p = 0.01 |
|  | Severe Day 0 - Moderate Day 0 | 10189.3 | 2045.6 | 6180.0 | 14198.6 | p < 0.001 |
| pH | Severe Day 7 - Day 0 | 0.0917 | 0.0117 | 0.07 | 0.11 | p < 0.001 |
|  | Moderate Day 7 - Day 0 | 0.1104 | 0.0197 | 0.07 | 0.15 | p < 0.001 |
|  | Severe Day 0 - Moderate Day 0 | 0 | 0.01 | -0.02 | 0.01 | p = 0.94 |
| Platelet count, x10^3^/$\mu$L | Severe Day 7 - Day 0 | 76.5 | 10.6 | 55.8 | 97.3 | p < 0.001 |
|  | Moderate Day 7 - Day 0 | 83.5 | 7.5 | 68.9 | 98.2 | p < 0.001 |
|  | Severe Day 0 - Moderate Day 0 | -65.4 | 6.3 | -77.7 | -53.2 | p < 0.001 |
| Potassium, mmol/L | Severe Day 7 - Day 0 | 0.0 | 0.1 | -0.1 | 0.2 | p = 0.75 |
|  | Moderate Day 7 - Day 0 | -0.1 | 0.0 | -0.2 | 0.0 | p = 0.04 |
|  | Severe Day 0 - Moderate Day 0 | -0.2 | 0.0 | -0.3 | -0.1 | p < 0.001 |
| Procalcitonin, ng/mL | Severe Day 7 - Day 0 | -15.2 | 5.4 | -25.8 | -4.7 | p = 0.005 |
|  | Moderate Day 7 - Day 0 | -6.6 | 2.5 | -11.4 | -1.7 | p = 0.008 |
|  | Severe Day 0 - Moderate Day 0 | 13.7 | 3.9 | 6.1 | 21.3 | p < 0.001 |
| Sodium, mmol/L | Severe Day 7 - Day 0 | 1.8 | 0.8 | 0.3 | 3.4 | p = 0.02 |
|  | Moderate Day 7 - Day 0 | 0.8 | 1.1 | -1.2 | 2.9 | p = 0.42 |
|  | Severe Day 0 - Moderate Day 0 | -1.2 | 0.3 | -1.7 | -0.7 | p < 0.001 |
| Troponin all types, ng/mL | Severe Day 7 - Day 0 | -0.71 | 0.28 | -1.25 | -0.16 | p = 0.01 |
|  | Moderate Day 7 - Day 0 | -0.47 | 0.17 | -0.81 | -0.14 | p = 0.006 |
|  | Severe Day 0 - Moderate Day 0 | 0.40 | 0.19 | 0.04 | 0.77 | p = 0.03 |
| White blood cell count,  x10^3^/$\mu$L | Severe Day 7 - Day 0 | -3.2 | 3.4 | -9.8 | 3.4 | p = 0.34 |
|  | Moderate Day 7 - Day 0 | -0.7 | 0.7 | -2.1 | 0.6 | p = 0.27 |
|  | Severe Day 0 - Moderate Day 0 | 2.8 | 0.9 | 1.0 | 4.5 | p = 0.002 |

Supplemental Table 4: Statistical comparison of the initial values and trends (from day 0 to day 7 of hospitalization) for laboratory results between moderate and severe pediatric subgroups. Day 7 and day 0 mean values not shown.

*Abbreviations:* ALT = alanine aminotransferase, AST = aspartate aminotransferase, BNP = brain natriuretic peptide, BUN = blood urea nitrogen, CI = confidence interval, IL-6 interleukin-6, NT-proBNP = N-terminal prohormone BNP.

| **Lab Test** | **Severity Category** | **Children with Lab Available,**  **n (%)** | **Adults with Lab Available,**  **n (%)** |
| --- | --- | --- | --- |
| ALT (SGPT), IU/L | Severe | 496 (72) | 20,992 (81) |
|  | Moderate | 2,243 (50) | 70,489 (78) |
| AST (SGOT), IU/L | Severe | 534 (78) | 22,405 (86) |
|  | Moderate | 2,333 (52) | 74,195 (83) |
| Albumin (g/dL) | Severe | 510 (74) | 20,388 (78) |
|  | Moderate | 2,021 (45) | 56,924 (63) |
| BNP (all forms), pg/mL | Severe | 301 (44) | 12,833 (49) |
|  | Moderate | 745 (16) | 33,214 (37) |
| BUN, mg/dL | Severe | 595 (87) | 23,427 (90) |
|  | Moderate | 2,835 (63) | 80,901 (90) |
| BUN/Creatinine ratio | Severe | 140 (20) | 8,318 (32) |
|  | Moderate | 927 (20) | 35,890 (40) |
| Bilirubin (Conjugated/Direct), mg/dL | Severe | 305 (45) | 12,975 (50) |
|  | Moderate | 811 (18) | 30,214 (34) |
| Bilirubin (total), mg/dL | Severe | 541 (79) | 22,444 (86) |
|  | Moderate | 2,350 (52) | 74,197 (83) |
| Bilirubin - (Unconjugated/Indirect),  mg/dL | Severe | 153 (22) | 4,394 (17) |
|  | Moderate | 389 (9) | 8,732 (10) |
| Chloride, mmol/L | Severe | 600 (88) | 23,385 (90) |
|  | Moderate | 2,843 (63) | 80,712 (90) |
| Creatinine, mg/dL | Severe | 594 (87) | 23,459 (90) |
|  | Moderate | 2,852 (63) | 81,067 (90) |
| D-Dimer, mg/L FEU | Severe | 260 (38) | 13,066 (50) |
|  | Moderate | 703 (16) | 40,886 (46) |
| Erythrocyte Sed. Rate, mm/hr | Severe | 307 (45) | 8,328 (32) |
|  | Moderate | 956 (21) | 18,797 (21) |
| Ferritin, ng/mL | Severe | 333 (49) | 15,073 (58) |
|  | Moderate | 847 (19) | 48,201 (54) |
| Fibrinogen | Severe | 375 (55) | 8,983 (35) |
|  | Moderate | 624 (14) | 11,720 (13) |
| Glucose, mg/dL | Severe | 599 (87) | 23,483 (90) |
|  | Moderate | 2,904 (64) | 81,232 (90) |
| Hemoglobin, g/dL | Severe | 598 (87) | 23,268 (89) |
|  | Moderate | 2,967 (66) | 81,892 (91) |
| IL-6, pg/mL | Severe | 66 (10) | 2,145 (8) |
|  | Moderate | 80 (2) | 4,098 (5) |
| Lactate Dehydrogenase (LDH), units/L | Severe | 248 (36) | 14,263 (55) |
|  | Moderate | 568 (13) | 42,752 (48) |
| Lactate, mM | Severe | 447 (65) | 17,377 (67) |
|  | Moderate | 808 (18) | 37,260 (41) |
| Lymphocytes (absolute), x10^3^/$\mu$L | Severe | 449 (66) | 19,106 (73) |
|  | Moderate | 2,379 (53) | 70,898 (79) |
| Lymphocytes (relative), % | Severe | 575 (84) | 21,412 (82) |
|  | Moderate | 2,725 (60) | 76,113 (85) |
| NT pro BNP, pg/mL | Severe | 163 (24) | 7,009 (27) |
|  | Moderate | 504 (11) | 21,233 (24) |
| Neutrophils (absolute), x10^3^/$\mu$L | Severe | 394 (58) | 19,510 (75) |
|  | Moderate | 2,107 (47) | 67,944 (76) |
| Neutrophils (relative), % | Severe | 407 (59) | 18,824 (72) |
|  | Moderate | 2,074 (46) | 69,612 (78) |
| Platelet count, x10^3^/$\mu$L | Severe | 487 (71) | 23,270 (89) |
|  | Moderate | 2,582 (57) | 81,524 (91) |
| Potassium, mmol/L | Severe | 597 (87) | 23371 (90) |
|  | Moderate | 2833 (63) | 80605 (90) |
| Procalcitonin, ng/mL | Severe | 157 (23) | 9982 (38) |
|  | Moderate | 543 (12) | 29025 (32) |
| Sodium, mmol/L | Severe | 600 (88) | 23393 (90) |
|  | Moderate | 2851 (63) | 80699 (90) |
| Troponin all types, ng/mL | Severe | 271 (40) | 12460 (48) |
|  | Moderate | 486 (11) | 26325 (29) |
| White blood cell count, x10^3^/$\mu$L | Severe | 480 (70) | 22982 (88) |
|  | Moderate | 2560 (57) | 80027 (89) |
| c-reactive protein CRP, mg/L | Severe | 408 (60) | 13653 (52) |
|  | Moderate | 1089 (24) | 33505 (37) |
| pH | Severe | 367 (54) | 15439 (59) |
|  | Moderate | 569 (13) | 18393 (20) |

Supplemental Table 5: Number and percent of children in the moderate (N = 4,528) and severe (N = 685) disease subgroups alongside the number and percent of adults in the moderate (N = 89,815) and severe (N = 26,033) disease subgroups who had a given lab test result available during their inpatient hospital encounter.

| **Category** | **MIS-C (N = 439), n (%)** | **Acute COVID-19 (N = 4,251), n(%)** |
| --- | --- | --- |
| Demographics | | |
| **Age** | | |
| Age < 1 year | <20 (0) | 773 (18) |
| Age 1-5 years | 100 (23) | 687 (16) |
| Age 5-12 years | 188 (43) | 680 (16) |
| Age 12-18 years | 143 (33) | 2111 (50) |
| **Sex** | | |
| Male | 259 (59) | 2066 (49) |
| Female | 180 (41) | 2185 (51) |
| **Race** | | |
| White | 181 (41) | 1875 (44) |
| Black | 143 (33) | 1032 (24) |
| Asian | <20 (0) | 78 (2) |
| **Ethnicity** | | |
| Hispanic | 111 (25) | 1168 (28) |
| Not Hispanic | 292 (67) | 2631 (62) |
| No Matching Ethnicity | 34 (8) | 431 (10) |
| Preselected Comorbidities | | |
| Obesity* | 121 (32) | 646 (30) |
| Asthma | 24 (6) | 425 (10) |
| Diabetes Mellitus (Type I or II) | <20 (0) | 129 (3) |
| Pediatric Complex Chronic Conditions | | |
| Any PCCC Category | 54 (12) | 1223 (29) |
| Cong Genetic PCCC | <20 (0) | 354 (8) |
| CV PCCC | 27 (6) | 424 (10) |
| GI PCCC | <20 (0) | 376 (9) |
| Heme Immune PCCC | <20 (0) | 405 (10) |
| Malignancy PCCC | <20 (0) | 204 (5) |
| Metabolic PCCC | <20 (0) | 369 (9) |
| Neonatal PCCC | <20 (0) | 170 (4) |
| Neuromuscular PCCC | <20 (0) | 403 (10) |
| Renal PCCC | <20 (0) | 237 (6) |
| Respiratory PCCC | <20 (0) | 231 (5) |
| COVID Testing | | |
| COVID +PCR/Ag | 156 (36) | 4251 (100) |
| COVID +Antibody | 369 (84) | <20 (0) |
| Laboratory Test | | |
| ESR (High) | 322 (73) | 403 (10) |
| Sodium (Low) | 280 (64) | 587 (14) |
| D-dimer (High) | 260 (59) | 356 (8) |
| LDH (High) | 251 (57) | 361 (9) |
| Albumin (Low) | 245 (56) | 334 (8) |
| CRP (High) | 232 (53) | 224 (5) |
| ALC (Low) | 209 (48) | 671 (16) |
| NT Pro BNP (High) | 202 (46) | 171 (4) |
| Ferritin (High) | 183 (42) | 149 (4) |
| Platelet Count (Low) | 166 (38) | 379 (9) |
| Procalcitonin (High) | 153 (35) | 121 (3) |
| ALT (High) | 149 (34) | 371 (9) |
| BNP (High) | 142 (32) | 42 (1) |
| WBC (High) | 138 (31) | 480 (11) |
| WBC (Low) | 82 (19) | 443 (10) |
| IL6 (High) | 63 (14) | 47 (1) |
| Hgb (Low) | 44 (10) | 217 (5) |
| Fibrinogen (Low) | 20 (5) | 49 (1) |
| ANC (Low) | <20 (0) | 69 (2) |
| Clinical Outcomes | | |
| Invasive Mechanical Ventilation | 52 (12) | 261 (6) |
| ECMO | <20 (0) | <20 (0) |
| Vasopressor / Inotropic Support | 135 (31) | 242 (6) |
| Deaths | <20 (0) | 50 (1) |

Supplemental Table 6: Number and percent of children in the MIS-C and acute COVID-19 subgroups with a given demographic characteristic, pre-existing comorbidity, abnormal lab value during hospitalization, or clinical outcome.

*The percent of children with obesity was calculated by dividing the number of children ≥2-years-old who had a BMI for age and sex ≥95th percentile in that subgroup (N = 121 for MIS-C and N = 646 for acute COVID-19), by the number of children in that subgroup who were ≥2 years old and had a BMI measurement available (N = 373 for MIS-C and N = 2,133 for acute COVID-19).

**Supplemental Figures**

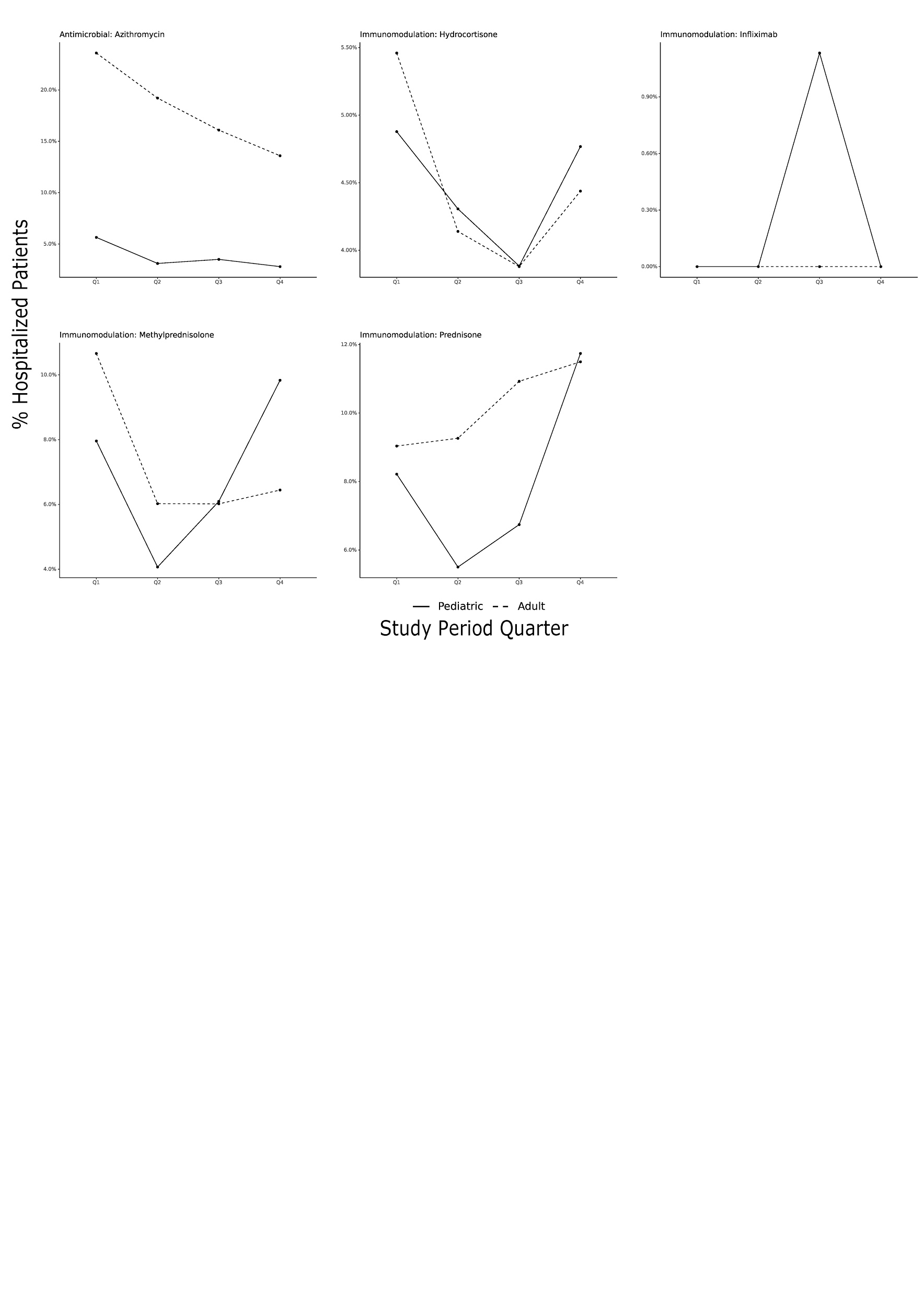

Supplemental Figure 1: Additional antimicrobial and immunomodulatory medication usage trends among hospitalized children and adults by study period quarter. Q1 = Apr 2020 - Jun 2020, Q2 = Jul 2020 - Sep 2020, Q3 = Oct 2020 - Dec 2020, and Q4 = Jan 2021 - Mar 2021.

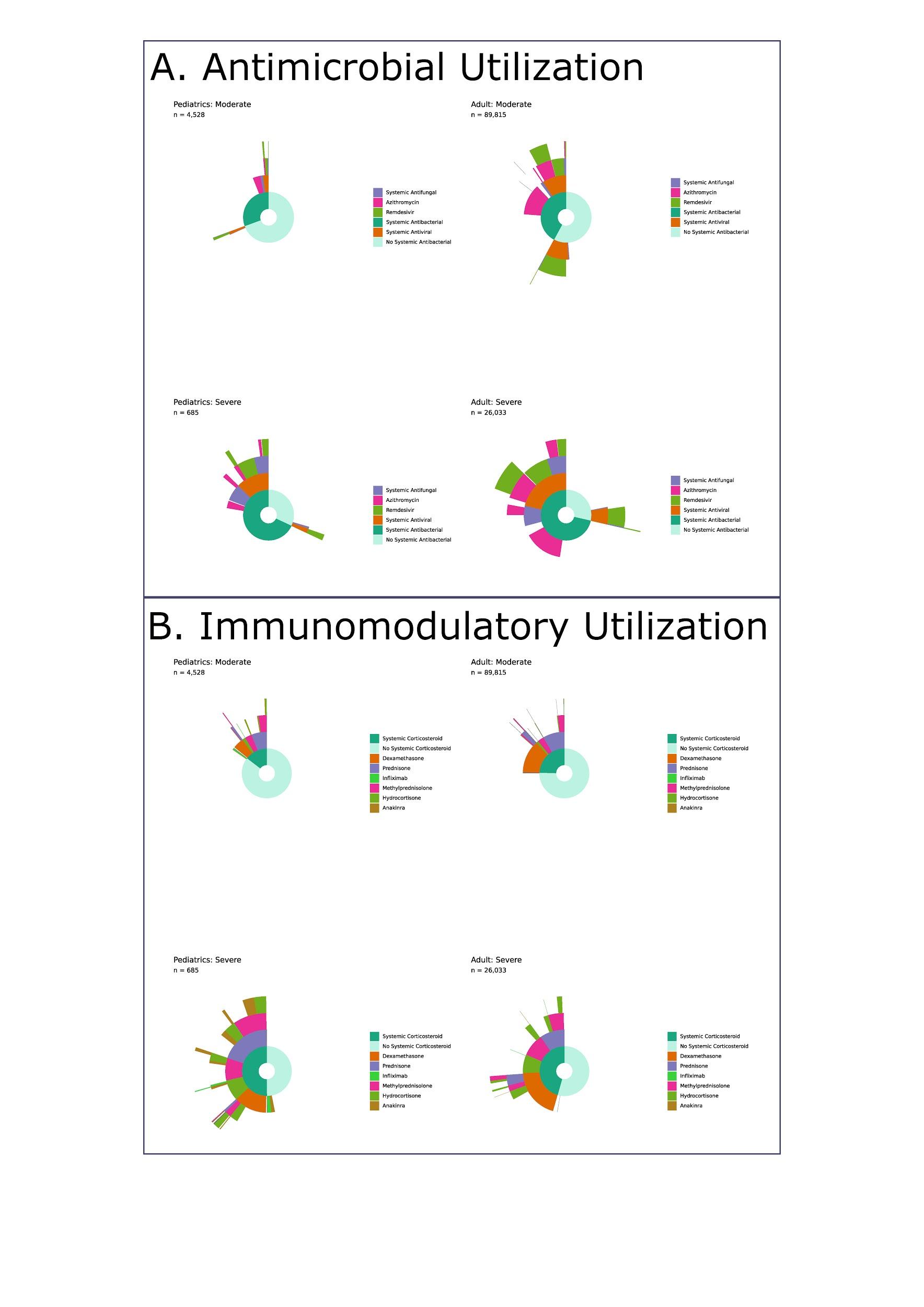

Supplemental Figure 2: Starburst plot of (a) antimicrobial and (b) immunomodulatory treatments in hospitalized children and adults by severity category.

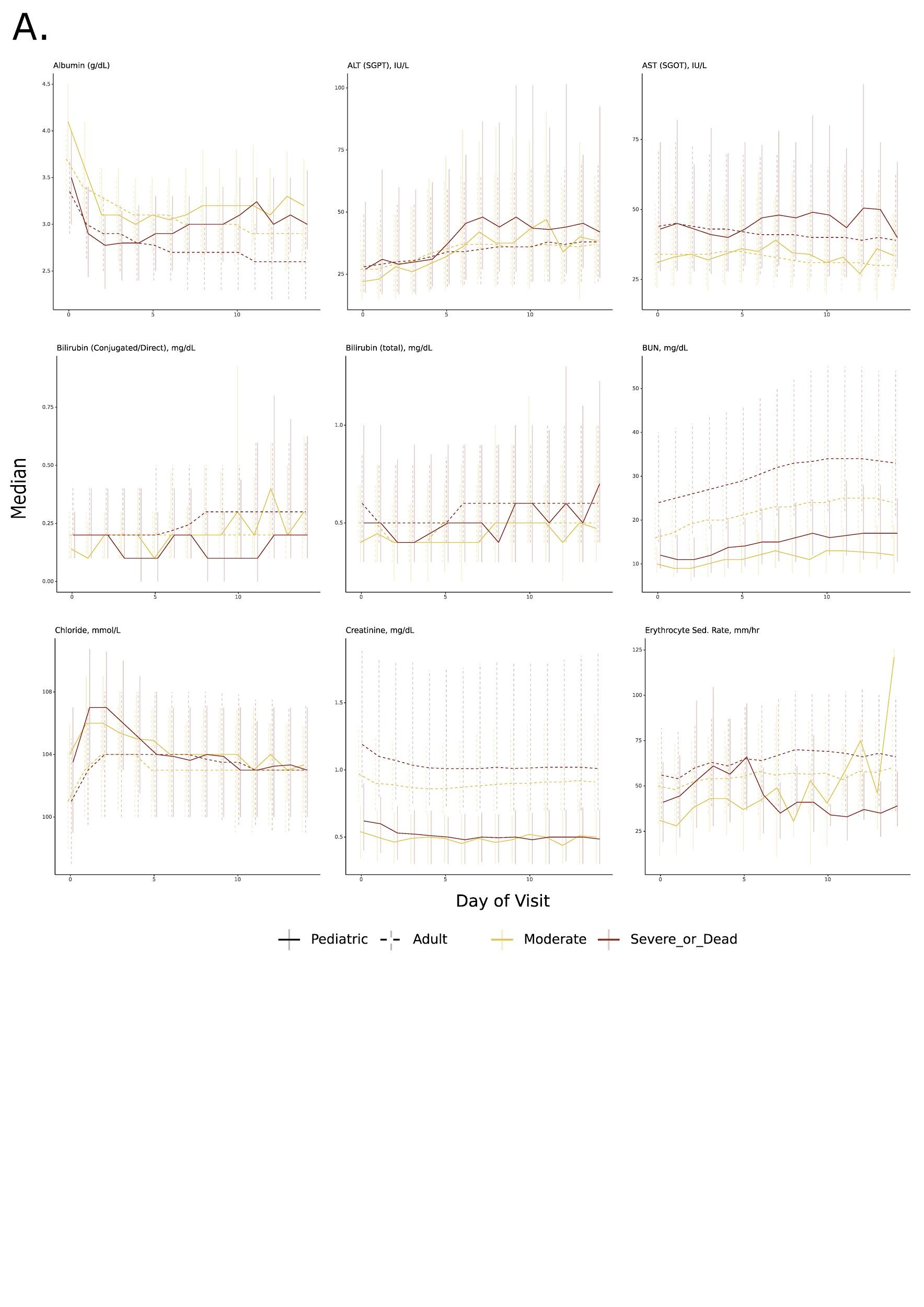

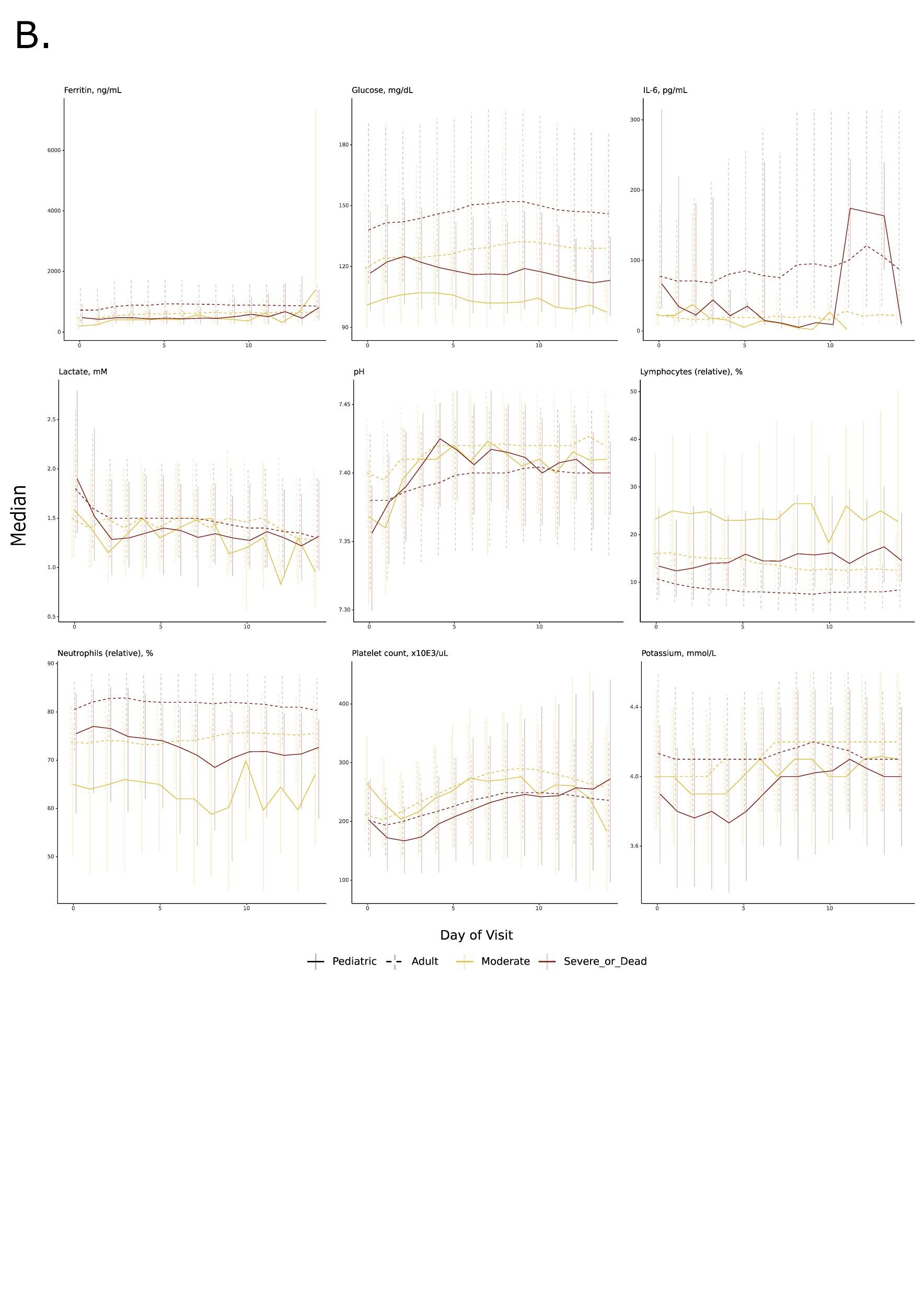

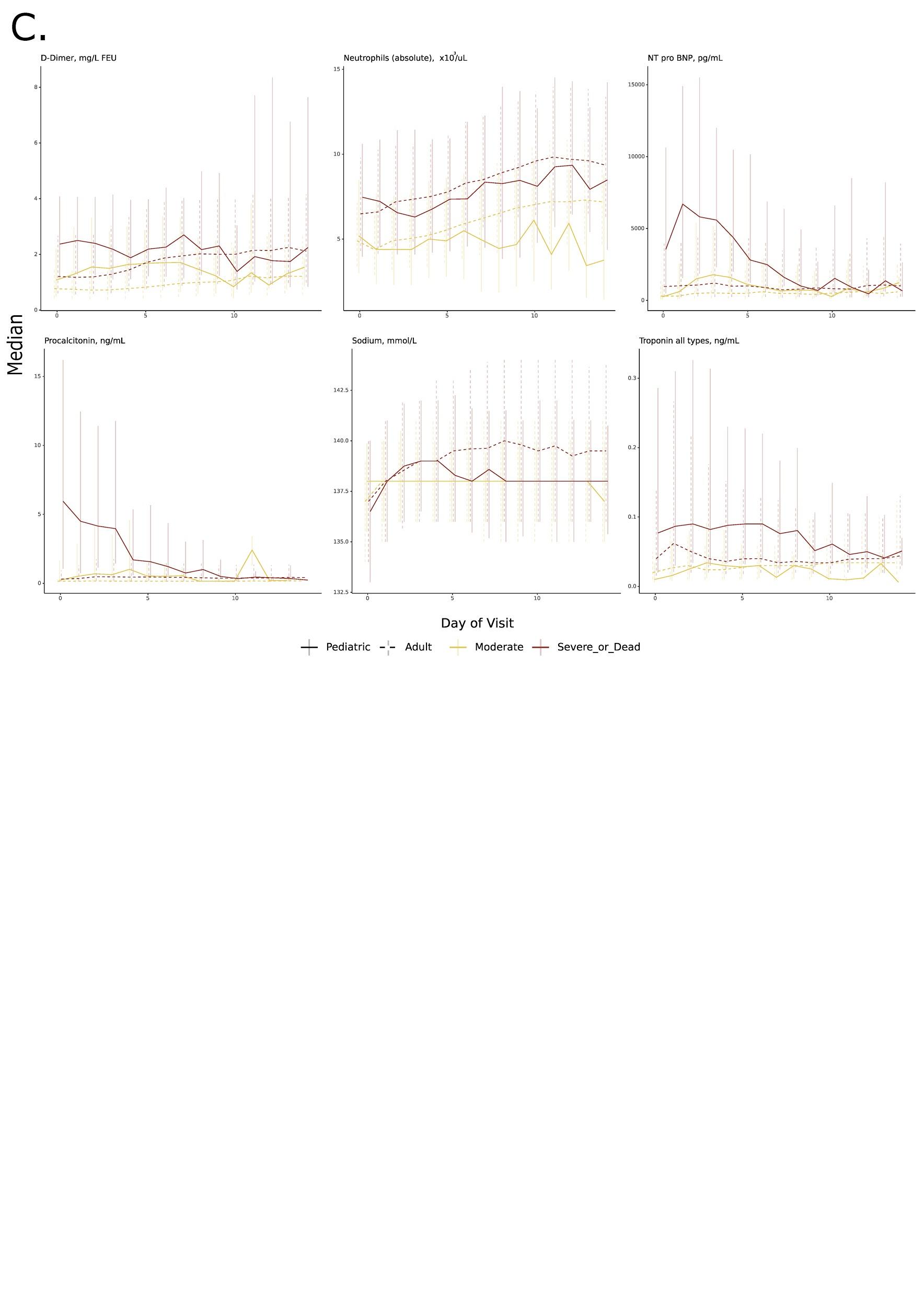

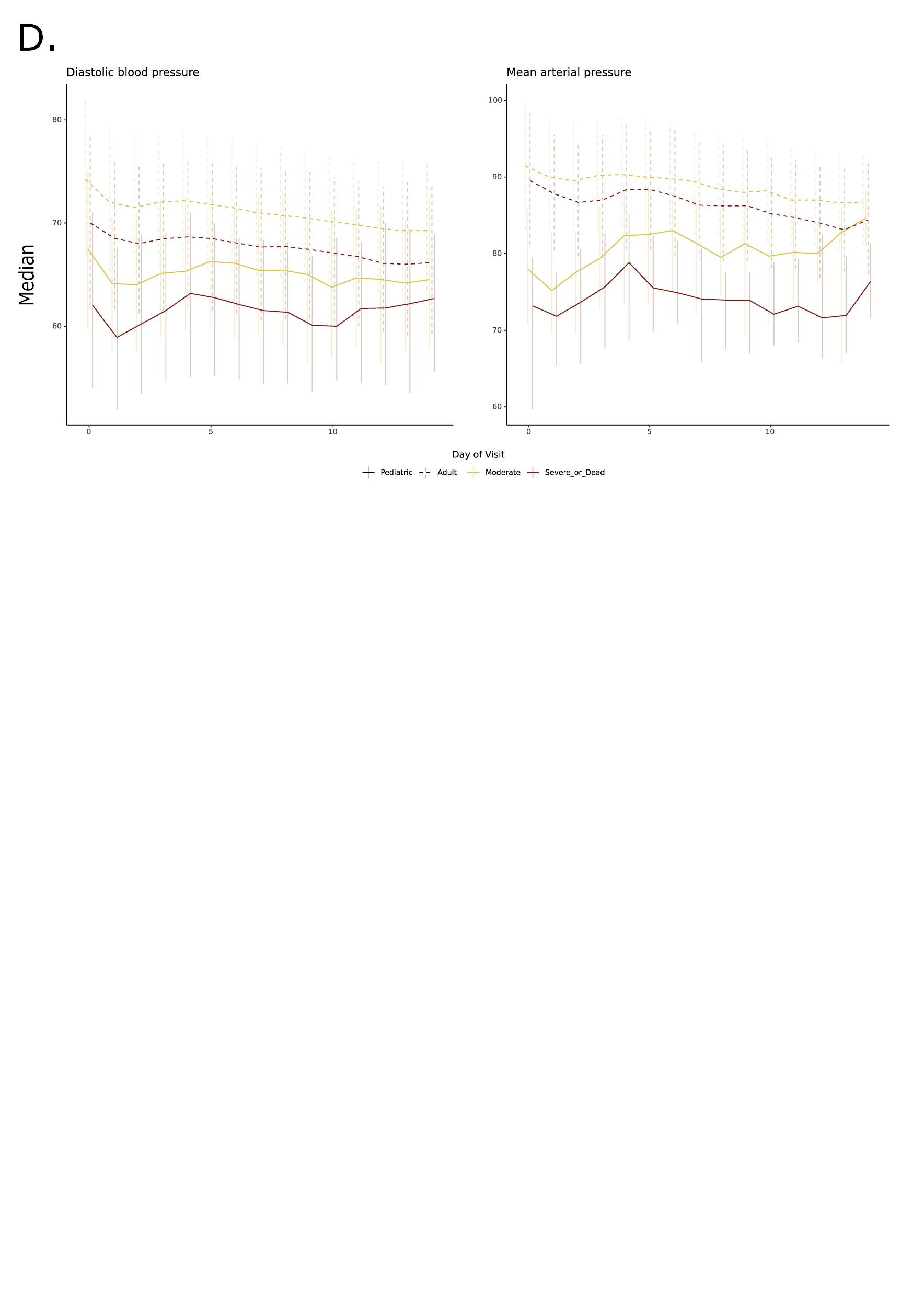

Supplemental Figure 3: Additional selected laboratory (a-c) and vital sign (d) median value trends by day of hospitalization for each pediatric and adult illness severity category.

*Abbreviations:* ALT = alanine aminotransferase, AST = aspartate aminotransferase, BUN = blood urea nitrogen, IL-6 = interleukin-6
